# Modeling the Effect of Lockdown Timing as a COVID-19 Control Measure in Countries with Differing Social Contacts

**DOI:** 10.1101/2020.11.14.20231886

**Authors:** Tamer Oraby, Michael G Tyshenko, Jose Campo Maldonado, Kristina Vatcheva, Susie Elsaadany, Walid Q Alali, Joseph C Longenecker, Mustafa Al-Zoughool

## Abstract

The application, timing, and duration of lockdown strategies during a pandemic remain poorly quantified with regards to expected public health outcomes. Previous projection models have reached conflicting conclusions about the effect of complete lockdowns on COVID-19 outcomes. We developed a stochastic continuous-time Markov chain (CTMC) model with eight states including the environment (SEAMHQRD-V), and derived a formula for the basic reproduction number, R_0_, for that model. Applying the *R*_0_ formula as a function in previously-published social contact matrices from 152 countries, we produced the distribution and four categories of possible *R*_0_ for the 152 countries and chose one country from each quarter as a representative for four social contact categories (Canada, China, Mexico, and Niger). The model was then used to predict the effects of lockdown timing in those four categories through the representative countries. The analysis for the effect of a lockdown was performed without the influence of the other control measures, like social distancing and mask wearing, to quantify its absolute effect. Hypothetical lockdown timing was shown to be the critical parameter in ameliorating pandemic peak incidence. More importantly, we found that well-timed lockdowns can split the peak of hospitalizations into two smaller distant peaks while extending the overall pandemic duration. The timing of lockdowns reveals that a “tunneling” effect on incidence can be achieved to bypass the peak and prevent pandemic caseloads from exceeding hospital capacity.

## Introduction

A cluster of viral pneumonia cases led to identification of a new coronavirus disease 2019 (COVID-19) first reported in Wuhan, China on December 31, 2019. Subsequent reports of human transmission [1] and travel-related cases [2] seeded outbreaks in many other countries. The WHO declared a global pandemic, Phase 6 emergency on January 30, 2020 [3].

Different country responses to early identified travel-related cases included quarantines and contact tracing [4] to identify and isolate potentially infected individuals, as containment measures. As the outbreaks developed, countries increased diagnostic testing of individuals with COVID-19 risk factors, respiratory symptoms and influenza like illness. However, once widespread local community was confirmed, transmission was present, frequently followed by discrete acceleration events [5], it rapidly overwhelmed the ability of many public health departments to conduct effective contact tracing and that of the health care system to take care of patients with critical, severe, and moderately severe illness; In response, many jurisdictions adopted mitigation broader strategies to manage the outbreak and slow down the rate of transmissions within the country such as social distancing, quarantines and lockdowns.

The surge in COVID-19 cases during the global pandemic put substantial strain on hospitals and intensive care units in China and other countries [6]. Interventions in China showed that contact tracing with quarantine, social distancing, and lockdowns to isolate cities and regions with community transmission was effective. The interventions in China were encouraging for modulating and containing the COVID-19 outbreak.

Non-pharmaceutical interventions (NPIs) that limit contact between individuals are proven to be efficacious in reducing COVID-19 transmission [7]. Contact limiting strategies include school closures, workplace closures (e.g. work-from-home mandate), stay at home orders and restrictions (e.g. for individuals, regions or entire countries), preventing gatherings (e.g. cancellation of larger events and smaller meetings), limiting visitors to institutional settings (e.g. hospitals, long term care facilities and prisons), voluntary or involuntary quarantine of potentially exposed individuals, quarantine of buildings, regions or lockdowns of entire countries (e.g. stopping most border traffic and international air travel). Various intervention strategies to reduce transmission can be utilized and are viewed as temporary public health measures [8].

Limiting contact is a strategy that attempts to decrease both the frequency and duration of contacts which in turn reduces the basic reproduction number, R_0_, the average number of persons to whom one case transmits the disease during his/her incubation period. Studies on social contact estimated that schools and daycare centers were the most socially dense locations compared with offices and homes [9]. When school closures and work-from-home strategies are activated, the transmission dynamics shift to the within-households contacts. In this regard, family structures, country population density, country population demographics, and socioeconomics can affect the number of social contacts occurring within the home. In addition, there is a problem of increased contact between individuals house-to-house, which may warrant a complete lockdown within the home.

Modelling with data fitted to Wuhan’s lockdown in China revealed a positive effect reducing the contact rate through isolation and quarantine that decreased and delayed COVID-19 infections [10]. However, other research studies suggest ongoing uncertainty over whether lockdown measures are sufficient to control 2019-nCoV [11].

To better understand the effect of lockdown dynamics for duration and timing, we created a stochastic continuous-time Markov chain model to analyze different hypothetical lockdown scenarios for four countries (Canada, China, Mexico, and Niger). The countries were chosen for their variation in social contact rates and ordered by increasing contacts using a scale of differential contract rates based on *R*_0_.

## Methods and Model

### Model Description

We used an SEAMHQRD-V disease transmission model to depict the spread of SARS-CoV 2 (the cause of COVID-19) in the community, and within households. The model is constructed from a stochastic continuous-time Markov chain (CTMC) with eight states or compartments: susceptible (S), exposed (E), infected but asymptomatic (A), mildly infected and symptomatic (M), severely infected, symptomatic and hospitalized (H), detected and quarantined (Q), recovered (R), and dead (D) (Fig. 1). The equivalent number of infected persons represented by deposition of virus particles by infected persons (A+M) in the environment is denoted by V, with removal of virus from the environment by ρ. Compartments were split into three age groups: children (0–18 years), denoted by a (c) subscript, adults (19–64 years) denoted by an (a) subscript, and senior (65 years and more), denoted by an (s) subscript. The possible transitions of individuals between compartments are represented by arrows with rates given above the arrows in Fig. 1. The subpopulation sizes are denoted by *N*_*c*_, *N*_*a*_, and *N*_*s*_, respectively, and they add up to the total population size *N* which is assumed to be fixed. See the supplementary material (S1. Model Description) for full description and Table S3 for definition of model’s parameters. We used the methods introduced by Allen and van den Driessche [12] to derive *R*_0_ for the CTMC model by approximating it by a multi-type branching process [12], see supplementary material (S2. The Basic Reproduction Number R_0_ and Probability of Extinction).

**Figure 1.**
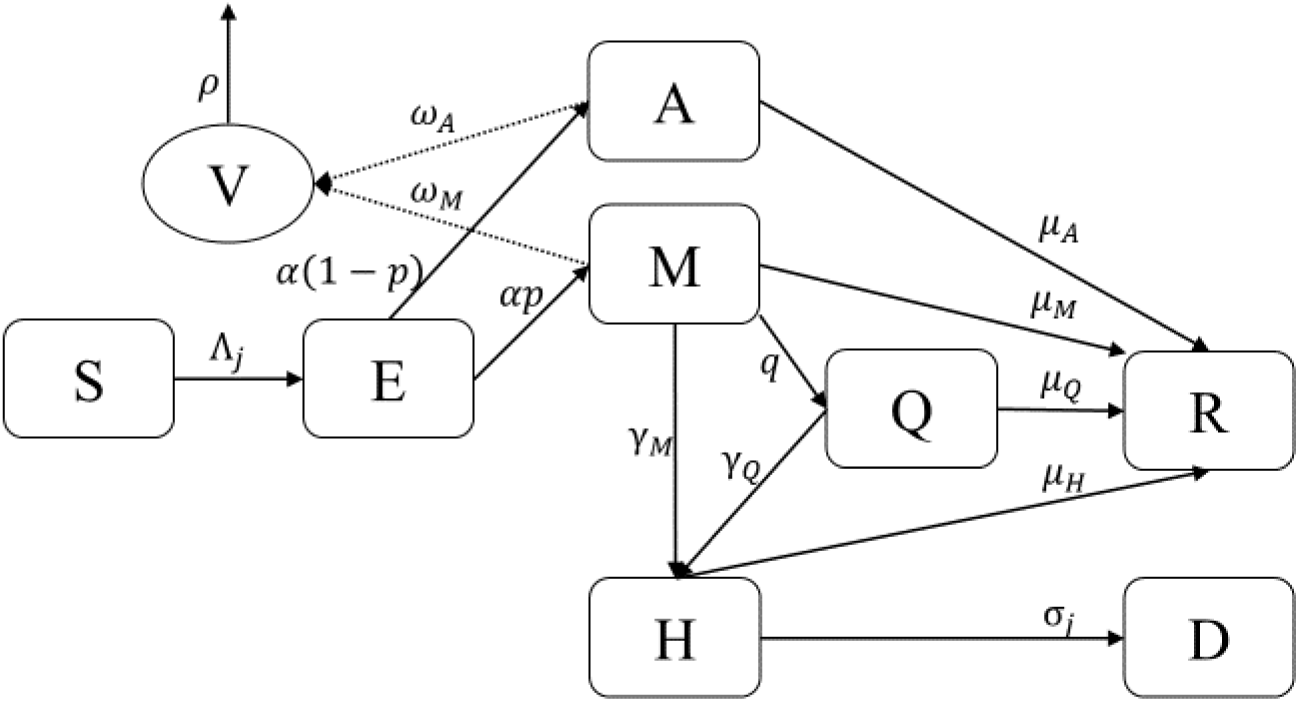
Schematic digraph of transitions of individuals between compartments in which transmission and transition rates are indicated over the arrows. See Table S3 for definition of model’s parameters. The force of infection *Λ*_*j*_ is given in equation (1), which depend on the environmental contact matrix *(C*^*V*^) and social contact matrices *(C)* for school, work, household, and other.

We used the CTMC to simulate different epidemiological measures and find their statistics. The first measure was the actual incidence, defined to be the proportion of the newly infected individuals to the population every day over the course of the epidemic. The second measure was the total attack rate, defined as the fraction of people that contract the disease in an at-risk population over the epidemic period. The third measure is hospital case load, defined to be the fraction of the population that is hospitalized for COVID-19 treatment at any given time and find its peak. We used the total population as the denominator for all of the measures so as to be able to compare between different counties with different population sizes.

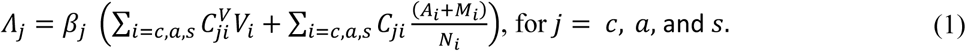

The social contact matrices used in the CTMC model were adapted from the study by Prem et. al. (2017) [13], which projected the data from population-based contact diaries in eight European countries from the POLYMOD study [14] to 144 other countries using a Bayesian hierarchical model that estimated age-and-location-specific contact patterns for the different countries. Applying household-level demographic and health survey data from the 152 countries to Markov chain Monte Carlo simulations, they produced five different types of social contact rates for various settings: work, school, home, other and all. The resulting social contact matrices were available for 5-year age increments from age 0 to 80.

To calculate the contact rates for our study for the assigned age groups of 0-18, 19-64 and 65+, we added up the contact rates of all columns (of the consequents) of the matrices (see Supplementary Material S1) representing age increments in each of these three age categories, and then averaged the totals across the rows (of the antecedent) for the corresponding compiled age groups. We assumed that environmental contact matrix is a proportion *r* of the all-contact matrix. The home contact matrix was normalized by the number of household members in each age group [15]. We obtained data about household sizes and population sizes classified by age groups in different countries from the United Nations, Department of Economic and Social Affairs, Population Dynamics [16].

We used the formula of *R*_0_ (calculated at β=3.5%) as a function in the contact matrices and demographics to produce the distribution and four categories of *R*_0_ for the 152 countries (Fig. S1) [13]; and chose one country from each quartile as a representative for each of four social contract categories. For this procedure, it was assumed that the probability of infection is the same in all situations and places. Quartiles of *R*_0_ split the countries into four groups (see S3. Countries Categorization for more details). We picked one country from each one of the four categories, that would also fall in a different continent: Canada, China, Mexico, and Niger; increasing from the lowest to the highest social contact category.

We used the tau-leap method [17] to simulate the stochastic CTMC model for 1,000 times. It is known that the size of the epidemic has a chance to be zero in CTMC models [18], which we exclude given that attack rates cannot, epidemiologically, have a value of zero and the COVID-19 virus has already a significant potential to spread between individuals. In all the simulations, the initial number of infected individuals was assumed to be one adult who is mildly infected. The list of parameters of the model, their description and values are shown in Table S3. Some of the parameters were found in the literature or guesstimated by experts and the rest of the parameters are found using calibration. We calibrated the model using the mean scenario that was estimated by [19] to be *R*_0_ = 6.47 since the model’s structure in [19] is very close to our model. That value was also very close to the mean value of the 152 values of *R*_0_ shown in Figure S1. We used that value for all the chosen countries to factor out the effect of the reproduction rate of secondary cases on the influence of the lockdown, thus allowing us to compare between the four countries.

To make a run-by-run comparison of the course of the epidemic with and without the lockdown, we simulated the model using a fixed random seed for each one of the 1,000 runs. In all those runs, we assumed that the only control measure is the complete lockdown starting before the peak of the actual incidence and for a specified period. The comparison of the effect of the start of the lockdown was done using two measures, which compute the relative reduction of measure X, RD(X), in *R*_0_, the total attack rate, and peak of hospitalization for the non-zero simulation runs with the following formula,

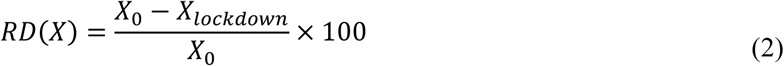

where *X*_0_ is the epidemiological quantity (*R*_0_, attack rate or peak of hospitalization) under no control measure and *X*_*lockdown*_ is the same measure with the lockdown. The mean, median and 2.5% and 97.5% percentiles of *RD* were calculated for the two measures. We also simulated the actual incidence and hospitalized normalized by the total population sizes for visual comparisons.

## Results

The basic reproduction number was derived (see Supplementary Material S2) and found to be proportional to the spectral radius ***(ρ)*** of a simple transformation of the contact matrix 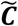 That is,

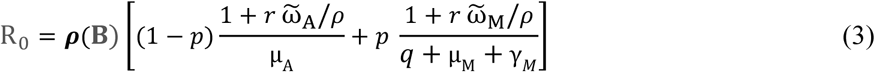

where

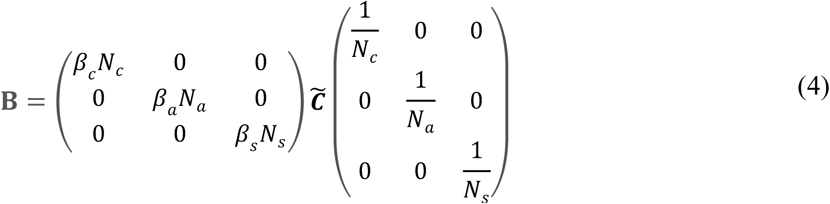

The matrix 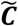 is the effective contact matrix based on social limitations (see Supplementary Material S1). The proportionality constant is dependent on the disease parameters, whereas the matrix **B** depends on social, demographic parameters as well as the transmission probabilities which are dependent on the strength of the virus and human culture and behavior.

The basic reproduction number *R*_0_ given in equation (3) shows direct dependence on the rates of viral environmental shedding and the fraction of contacts individuals make with the environment. Decreasing contact with the environment by use of personal protective equipment (PPE) and increasing environmental decontamination by frequent cleaning, disinfection, hand washing will result in a decrease in basic reproduction number. On the other hand, increasing rates of “removal” of both asymptomatic and mild cases, operationalized through contact tracing, isolation and quarantine, will lower the denominators of the term on the right side of equation (3) to a degree proportional to the spectral radius of the matrix ***B*** in equation (4). Strict adherence to social distancing and stopping insalubrious cultural habits by the three age groups can decrease the likelihood of disease transmission (through *β*’s) resulting in shrinking *R*_0_. Meanwhile, 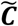 can be altered through mutual social limitations by individuals in each age group, like through partial or full lockdowns.

A full lockdown results in a reduction in the basic reproduction number *R*_0_ by more than 64% and up to 85% (Fig. 2). We can conclude that with using this model, household contacts and demographics are among the major factors contributing to *R*_0_.

**Figure 2.**
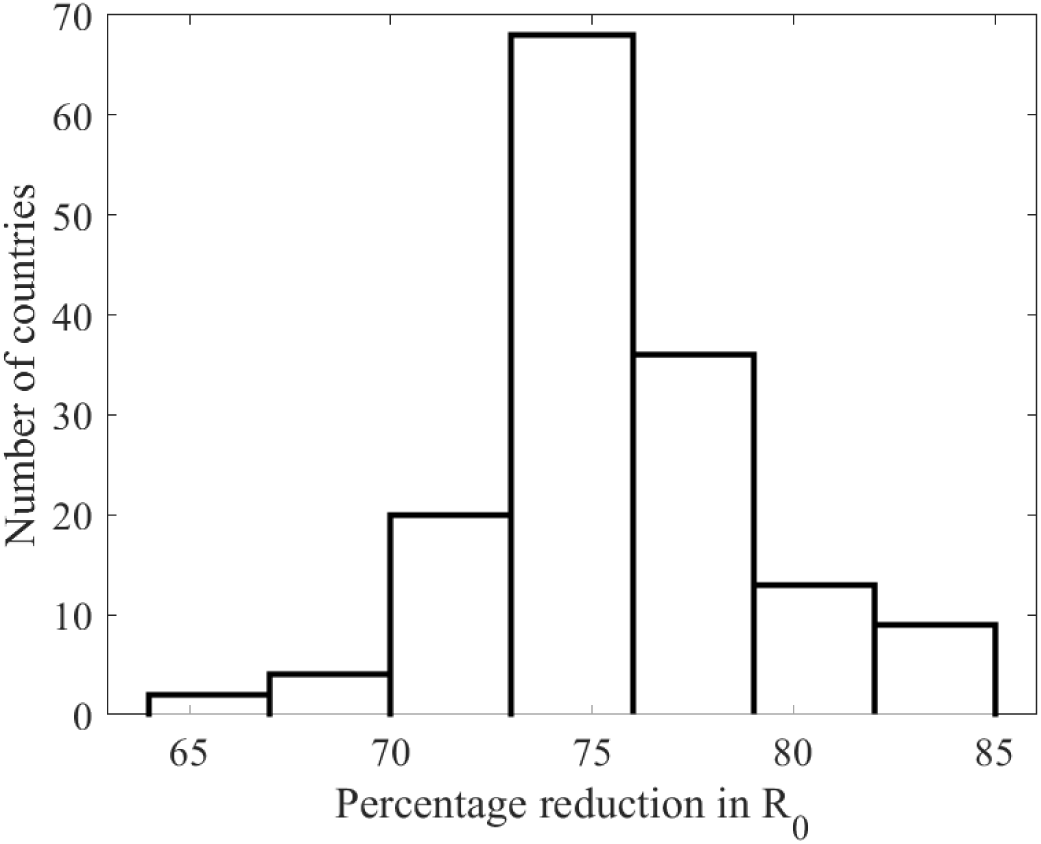
Histogram of percentage reduction in values of *R*_0_ for the 152 countries [13] calculated at β = 3.5% (see equation (2)). The percentage reduction of the four selected countries are as follows: Canada 82%, China 76%, Mexico 74%, and Niger 73%.

The start and length of lockdown affects attack rates, and hospital flux (Fig. 3 and 4) with different degrees. This is consistent with the various possible levels of reduction in the basic reproduction numbers as can be seen in Fig. 2. While the magnitude of relative reduction is not the same for the four selected counties, the consequences of timing and length of lockdown appears to be consistent. It appears that starting a complete lockdown will have its optimal reduction on the total attack rate if it starts 5 days before the peak of the actual incidence and lasting for 90 days. While starting a 90-days lockdown 30 days before the peak has a small relative reduction of the total attack rate (Fig. 2), it reduces the peak of the actual incidence (Fig S4). Shorter lockdowns seem to be of larger relative effect on the total attack rate if they start close enough to the peak of the actual incidence.

**Figure 3.**
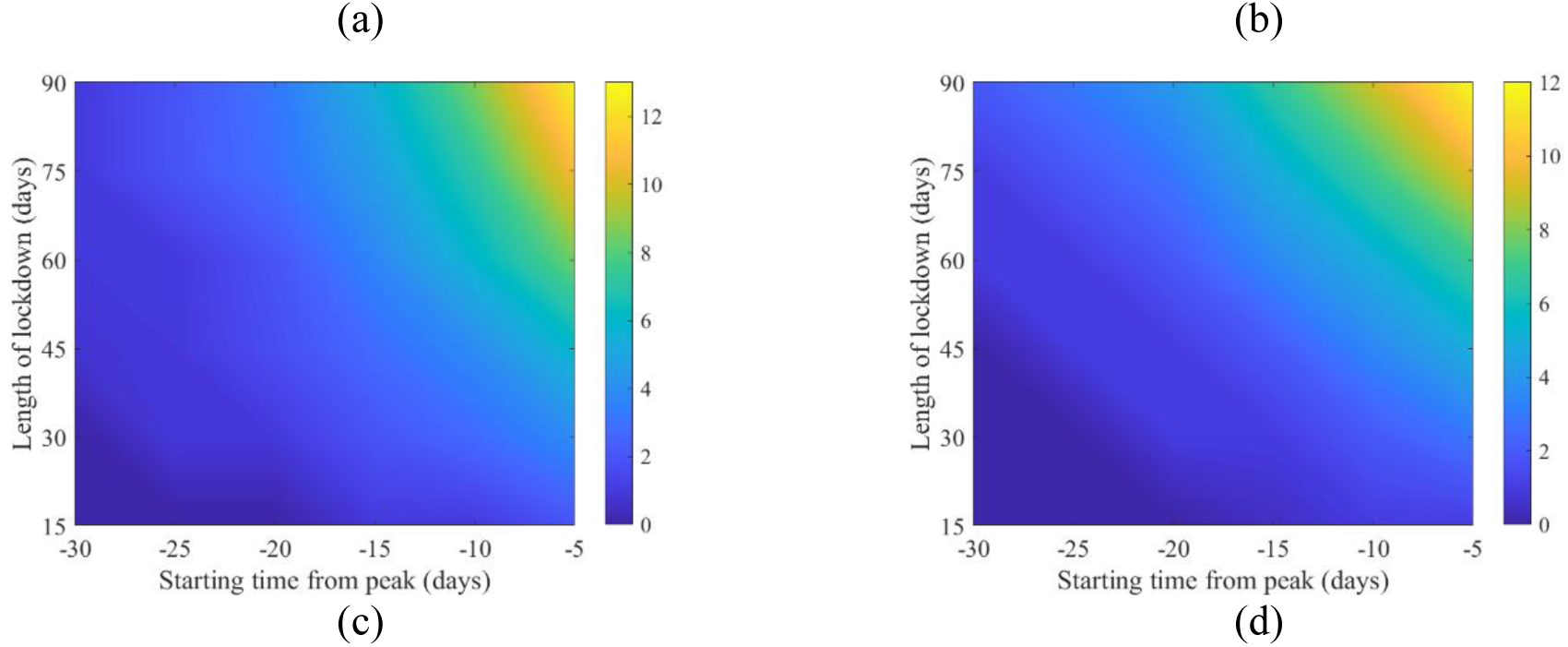
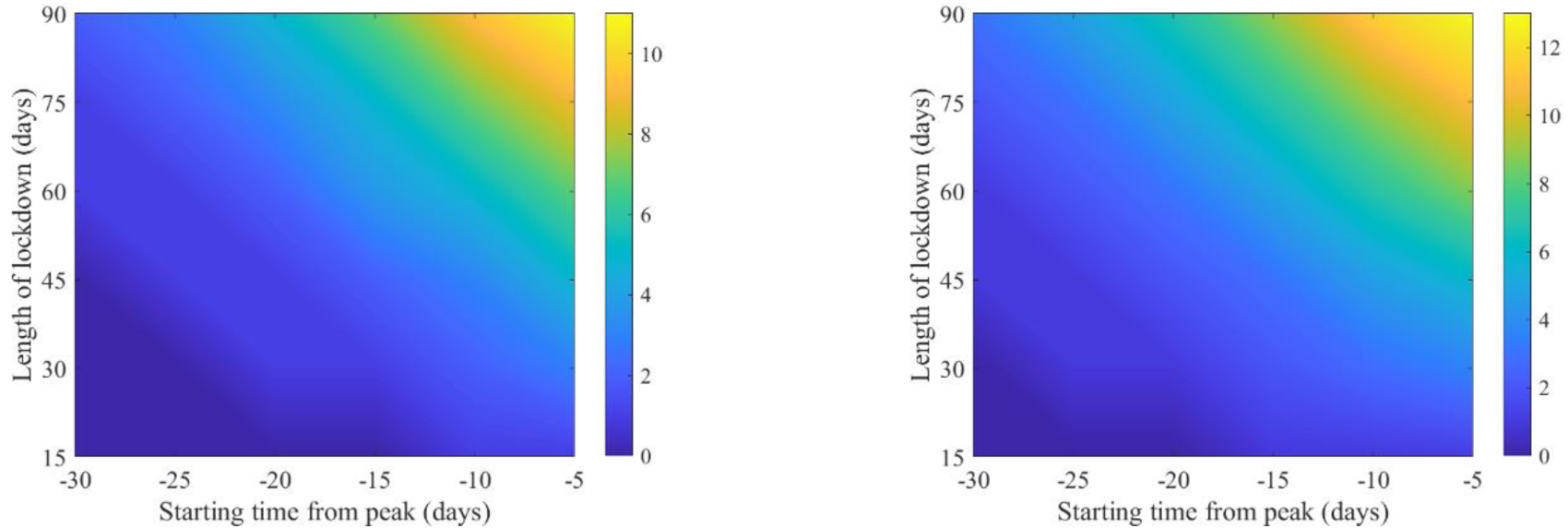
Mean of percentage relative reduction in COVID-19 total attack rates (see equation (2)) for (a) Canada, (b) China, (c) Mexico, and (d) Niger. They are calculated at *R*_0_ = 6.47, with initially one adult mild infection. Bars to the right of the figures are percentages.

**Figure 4.**
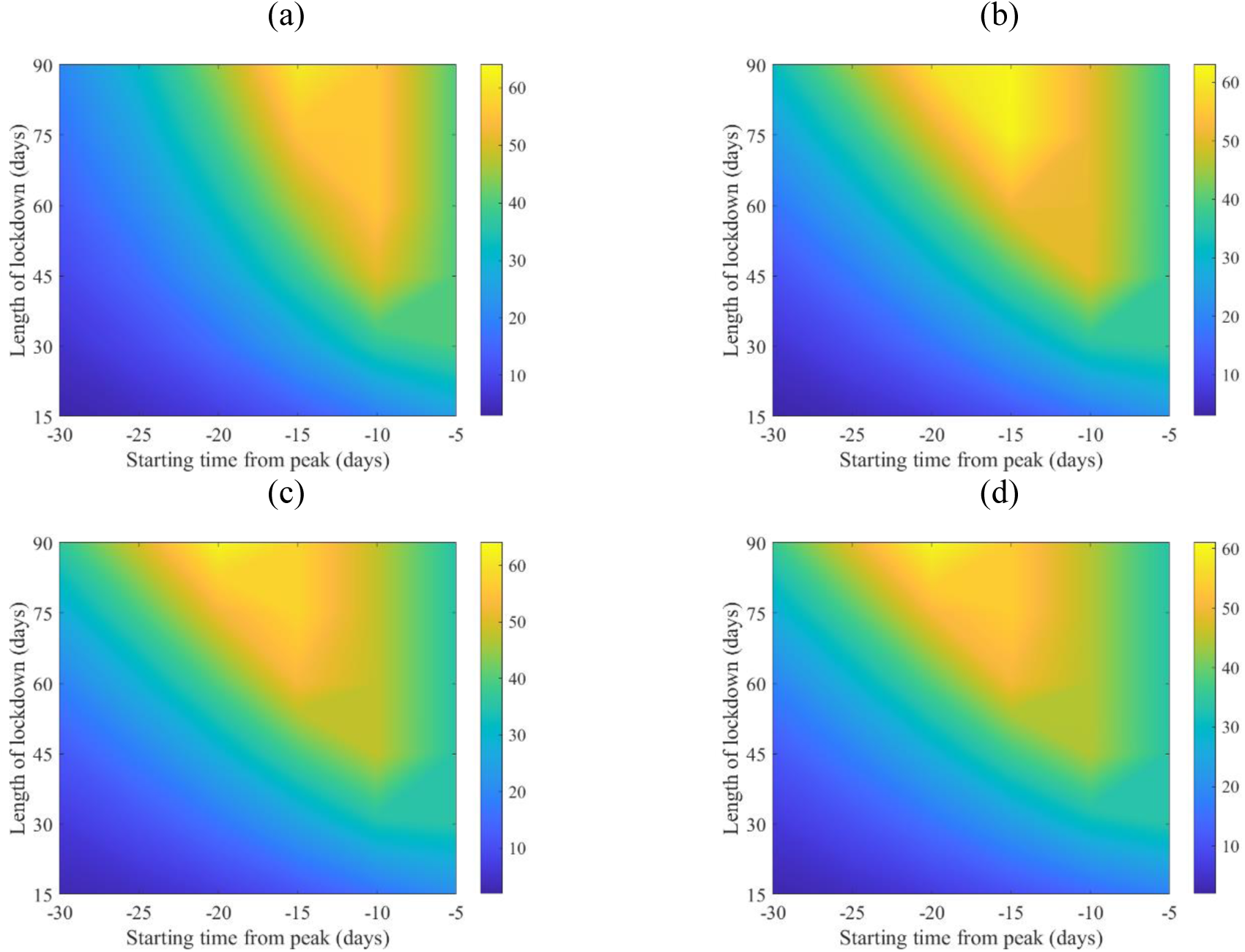
Mean of percentage relative reduction in peak of COVID-19 hospitalization (see equation (2)) for (a) Canada, (b) China, (c) Mexico, and (d) Niger. They are calculated at *R*_0_ = 6.47, with initially one adult mild infection. Bars to the right of the figures are percentages.

Lockdown, however, has its maximum effect on the hospital case load if it starts 15 days to 20 days prior to the peak of actual incidence, See Fig. 4. The timing is less consistent between the four countries but shows an overall qualitative resemblance. It appears that a shorter than a 90-days lockdown can achieve the goal of hospital case load reduction.

The optimality results are significant to a large degree as could be seen in the 95% quartiles interval of the percentage reduction in attack rate and hospitalization flux shown in Fig. S2 and S3.

While the location of the peak of the proportion of the actual incidence of COVID-19 cases to the total population is different for the four countries, the magnitudes of the peaks are very close, as shown in Fig. 5-8 (a), which might be an outcome of the contact matrices and demographics while keeping *R*_0_ constant. Starting the lockdown 15 days before the actual incidence’s peak results in a tunneling effect of the incidence curve as seen in the simulation runs and their average, Fig. 5-8 (b) (see also Fig. S4), which resulted in a decrease in the magnitude of the attack rates. The tunneling effect appears as a theoretical solution in environmental Kuznets curves of pollution emission [20]. Here it also results in dividing the flux of cases arriving at hospitals into two distinct, smaller peaks which would allow hospitals to deal with a smaller initial peak before restocking for the second smaller peak, Fig. 5-8 (c) and (d). Dividing the peak of hospitalization into two smaller peaks creates a more manageable outbreak scenario. Thus, hospitals can “divide and conquer” the expected larger peak of cases with a well-timed lockdown. The benefit of not exceeding hospital capacity is decreased mortality (not explicitly modeled here).

**Figure 5.**
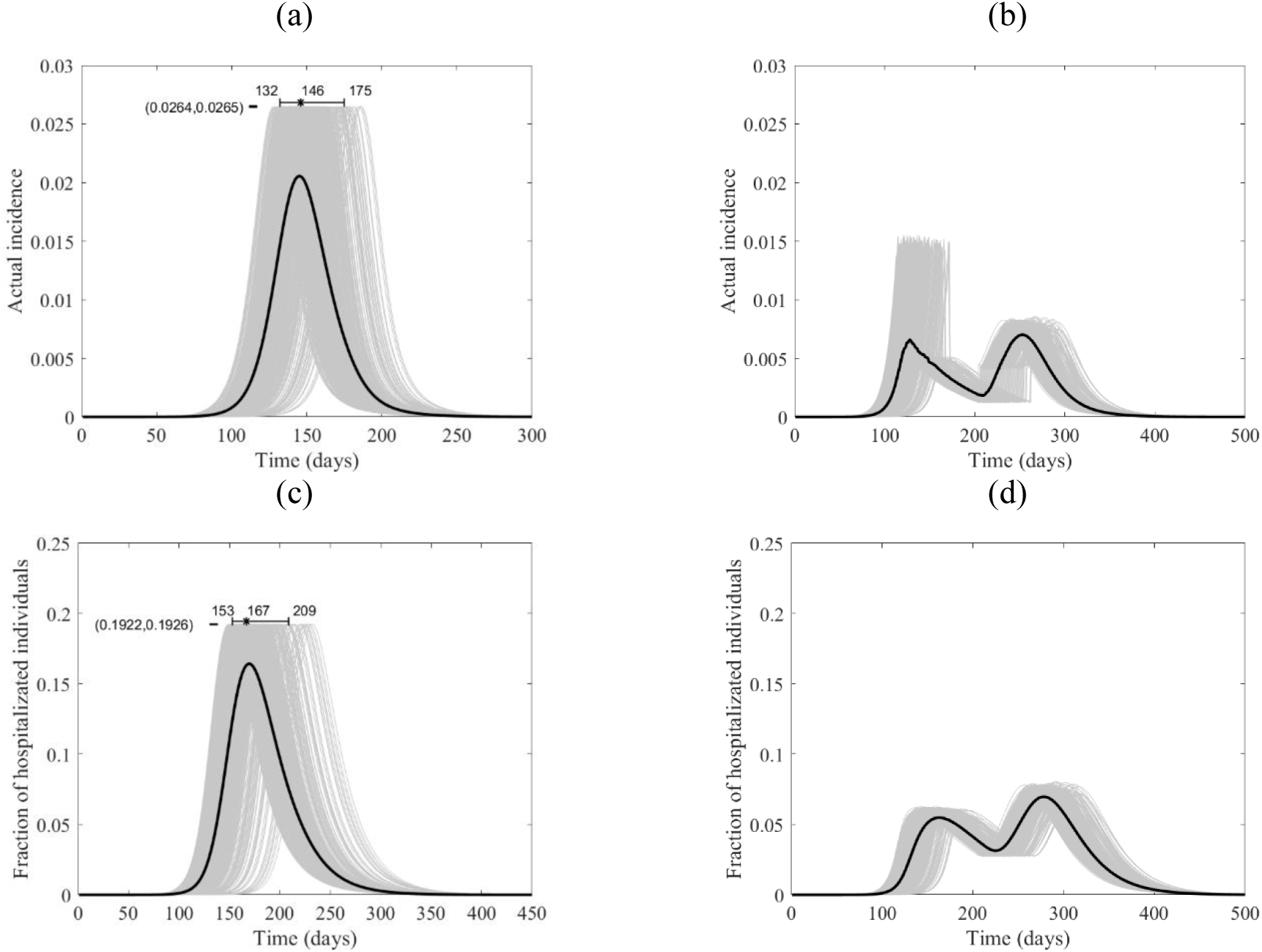
The course of the actual incidence (a) and (b), and fraction of hospitalized COVID-19 infected individuals (c) and (d) in Canada with no control measure (left panel) and with starting lockdown (right panel) of 15 days before the peak and that lasts for 90 days. They are calculated at *R*_0_ = 6.47, with initially one adult mild infection. The grey curves are resulting from the stochastic model simulations and the black curve is the mean of those grey curves. They are all normalized by the population size.

**Figure 6.**
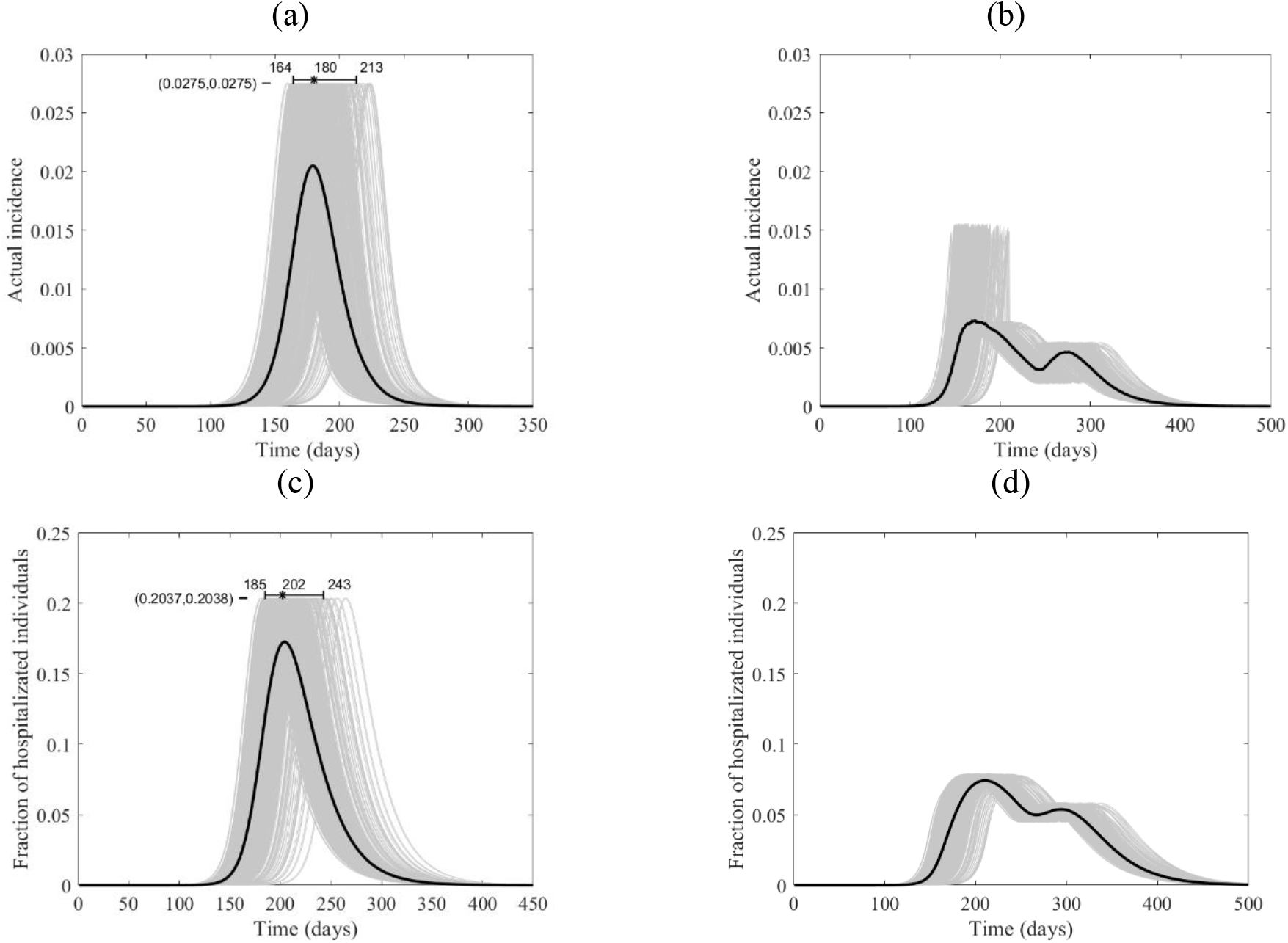
The course of the actual incidence (a) and (b), and fraction of hospitalized COVID-19 infected individuals (c) and (d) in China with no control measure (left panel) and with starting lockdown (right panel) of 15 days before the peak and that lasts for 90 days. They are calculated at *R*_0_ = 6.47, with initially one adult mild infection. The grey curves are resulting from the stochastic model simulations and the black curve is the mean of those grey curves. They are all normalized by the population size.

**Figure 7.**
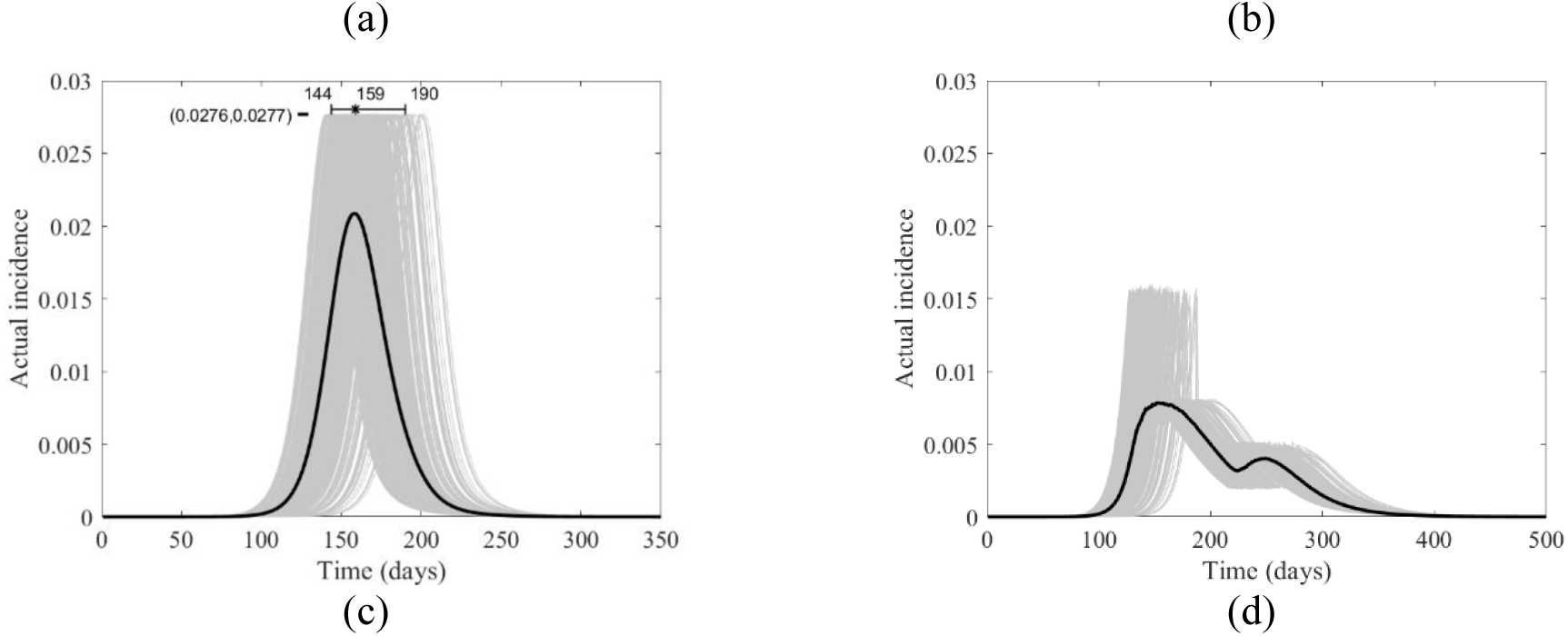
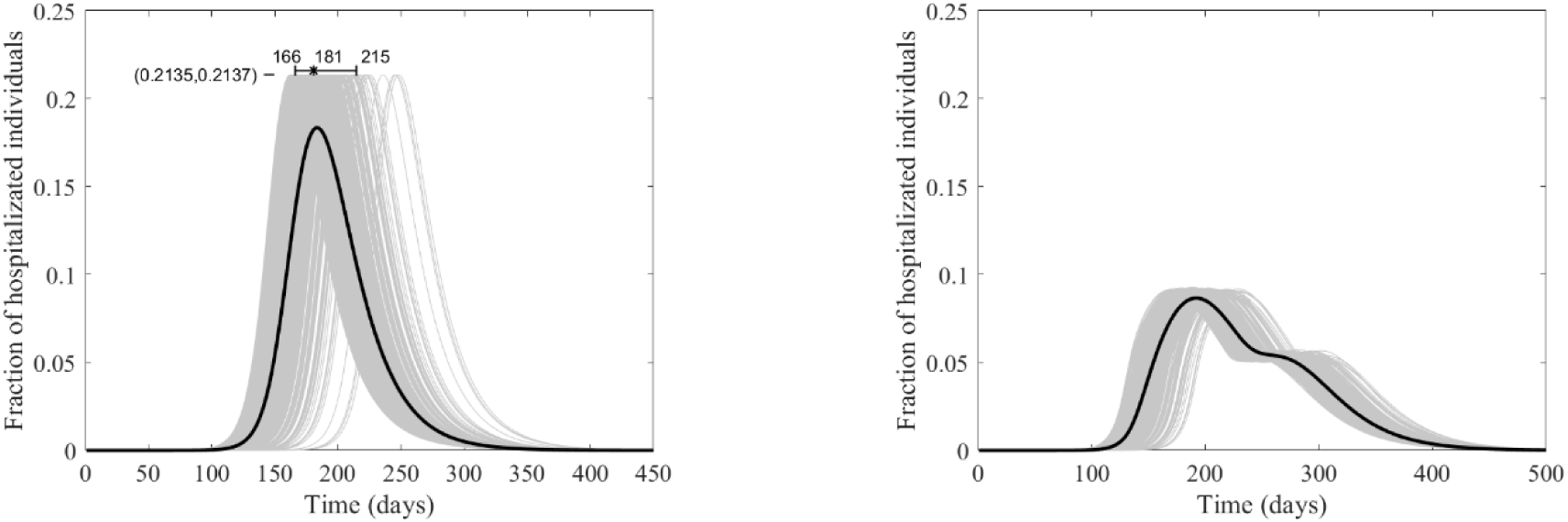
The course of the actual incidence (a) and (b), and fraction of hospitalized COVID-19 infected individuals (c) and (d) in Mexico with no control measure (left panel) and with starting lockdown (right panel) of 15 days before the peak and that lasts for 90 days. They are calculated at *R*_0_ = 6.47, with initially one adult mild infection. The grey curves are resulting from the stochastic model simulations and the black curve is the mean of those grey curves. They are all normalized by the population size.

**Figure 8.**
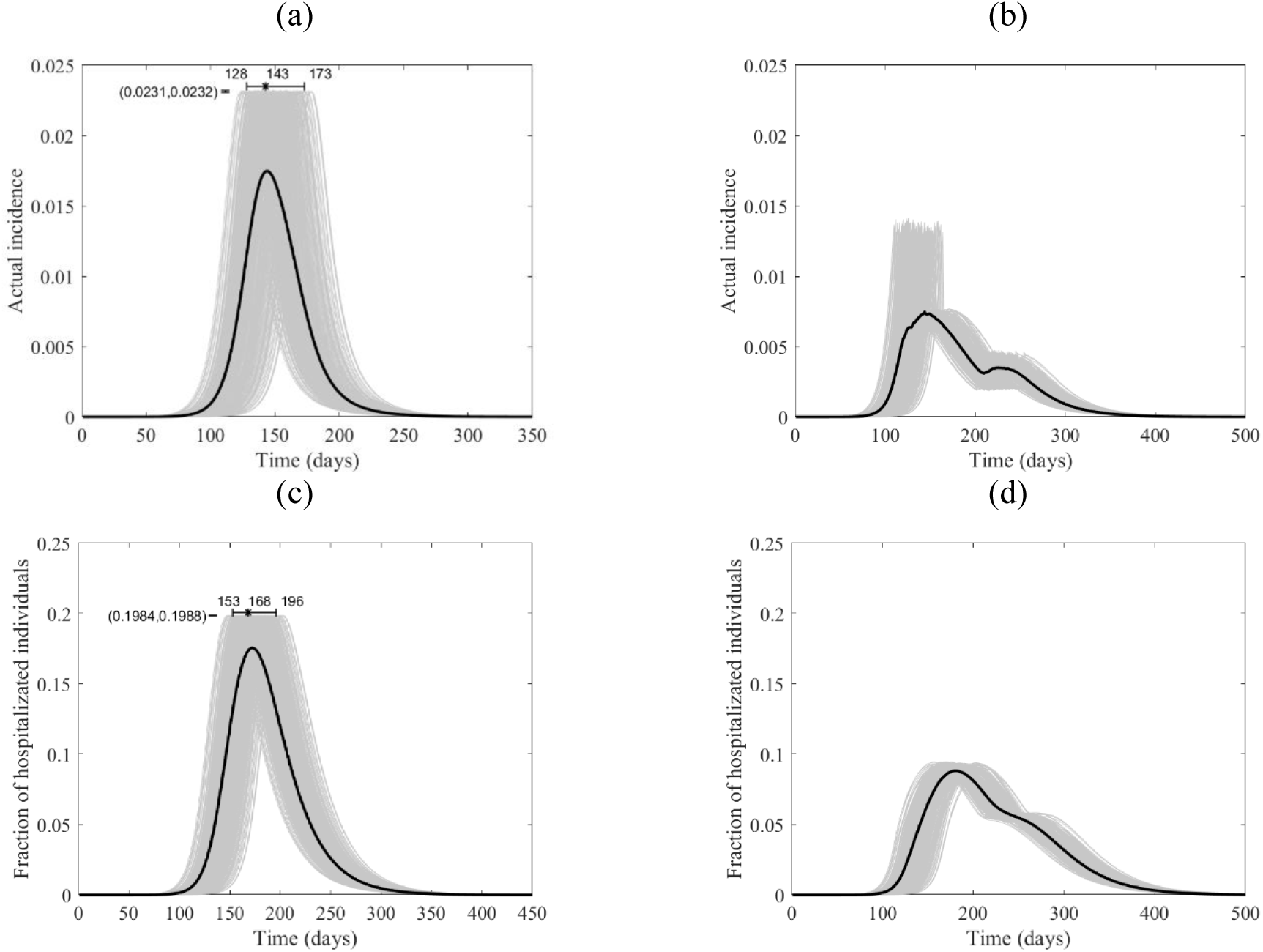
The course of the actual incidence of COVID-19 (a) and (b), and fraction of hospitalized infected individuals (c) and (d) in Niger with no control measure (left panel) and with starting lockdown (right panel) of 15 days before the peak and that lasts for 90 days. They are calculated at *R*_0_ = 6.47, with initially one adult mild infection. The grey curves are resulting from the stochastic model simulations and the black curve is the mean of those grey curves. They are all normalized by the population size.

While the effect of the length of the lockdown on the two measures is expected, the effect of its start is subtle. In Fig. 9, we can see some transitions in the hospitalizations based on the decision to start the lockdown according to the four timing options 20, 15, 10, and 5 days before the actual incidence peaks. The model output shows the 15-days before the peak lockdown option to be the most effective for Canada and China while the 20-days option seems best for Mexico and Niger.

**Figure 9.**
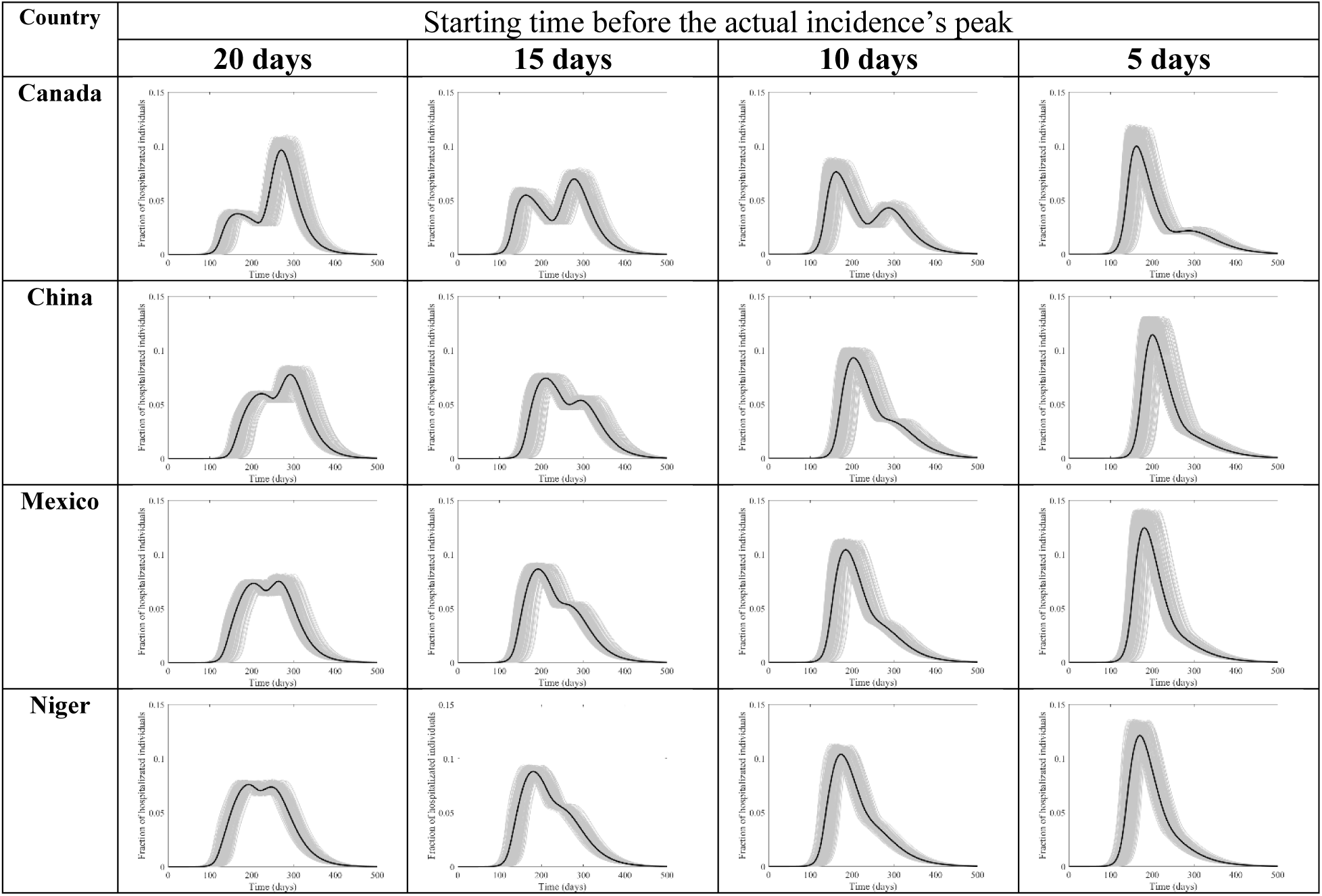
Hospitalization flux (proportion of COVD-19 cases requiring hospitalization) for Canada, China, Mexico, and Niger at four different times (days) of starting the lockdown before the peak. They are calculated at *R*_0_ = 6.47, with initial one adult mild infection. The grey curves are resulting from the stochastic model simulations and the black curve is the mean of those grey curves. They are all normalized by the population size.

Similarly, the actual incidence show transitions (Fig. S4) with the maximum effect on the peak happening when the lockdown starts 15-20 days before the peak of the actual incidence. Simulations of the total attack rates show reductions consistent with the results (Fig. S5-8)

## Discussion

Continuous time Markov chain models are regularly used to model transmission of diseases. To date most of the work done to model COVID-19 has used deterministic modelling which gives an approximation of the mean of the stochastic epidemic curves [12, 21-23]. However, the deterministic curves will miss the likely timing of the peak of incidence since averaging over values does not imply averaging over time. The outputs of our carefully crafted simulations of the (stochastic) CTMC model demonstrate that the timing of the lockdown relative to the epidemic peak is a key factor in controlling COVID-19 and prevent hospital systems from becoming overwhelmed.

According to this CTMC model, countries with different social contact rates revealed that the optimal starting time to decrease the total attack rate occurs when the lockdown begins about 5 days before the actual peak of the epidemic, which is the peak of incidence. Benefits from lockdown in terms of relative reduction in COVID-19 hospitalizations are also observed around 15-20 days before the epidemic peak. This provides a limited window for public health decision-makers to mobilize and take full advantage of lockdown as an NPI. Lockdowns appear to have a maximum effect if they start close to the actual peak of incidence and last for three months.

Timing the start of a complete lockdown 30 days or more prior to the epidemic peak will have little appreciable effect on reducing either the total attack rate or the peak of hospitalizations (as shown in Fig. 3 and 4). Accurate knowledge of the actual peak of incidence, not just of the reported cases may be required for optimal control of the epidemic using lockdown as an intervention, but obtaining a realistic timing of the peak and the actual incidence seem unattainable given the uncertainties involving the COVID-19 pandemic.

Starting the lockdown too early or too late can miss the chance of optimal benefit in controlling the disease. The output of this model illustrates that likely there is an optimal window to start the lockdown and provide maximum benefits for COVID-19 incidence and hospitalizations relative to the width of the 95% quartile interval of the location of the peak. Interventions must take into consideration repercussions such as major economic impacts, mental health consequences, and increased morbidity and mortality from non-COVID-19 diseases [24]. Poor timing will result in a wasted lockdown effort with little impact on the outbreak while incurring economic losses and psychological tolls to the public and healthcare workers during extended isolation including response and lockdown fatigue [25].

While the decision of when to begin a lockdown will vary from one country to another, based on their specific outbreak context, the knowledge of the location of the actual peaks makes a difference when comparing the difference between starting at 5 days and 15 days before the peak. Knowledge about delays in testing and reporting COVID-19 cases as well as accurate estimates of the epidemic are required to make evidence-based decisions.

When faced with COVID-19, countries have used multiple NPIs concurrently in various combinations and timings including lockdowns to reduce *R*_0_ [26]. Curbing *R*_0_ (see Equation 2) can be achieved by decreasing the probability of transmission via social distance and changes in cultural norms. Limiting contact, frequent cleaning and environmental disinfection as well as wearing masks and face coverings can also result in a linear decrease in *R*_0_.

Past studies showed that the strongest approach to limit social contacts is achieved through a partial or complete lockdown; e.g. China used aggressive city and regional lockdowns to prevent transmission from symptomatic and asymptomatic cases, thus flattening the epidemic curve and pushing the peak further into the future. Use of strict lockdowns along with other NPIs allow healthcare systems to treat a more manageable case load and gain time to plan, educate the public optimize the public health response while advancing efforts for vaccine development [27].

We modelled hypothetical lockdowns four weeks longer than those used in Wuhan and other Chinese cities without other NPIs, revealing that the timing of lockdowns is critical for its effectiveness in reducing actual incidence [28-29]. Our modeling shows the benefit of an accurately timed lockdown which results in a tunneling effect and can provide relief by avoiding overwhelming the hospitals, the public health response capabilities and the health care system. However, the lockdown, while reducing case load, can be expected to extend the duration of the epidemic. Extending its duration could result in unforeseen outcomes in healthcare such as higher immediate healthcare costs (e.g. extended need for personal protective equipment, testing kits, laboratory diagnostics, and increased intense hospital cleaning), decreased utilization of healthcare services for other diseases resulting in worse outcomes and increased mortality (e.g., reduced cancer screening or treatment for heart disease or diabetes), increased incidence of psychological outcomes (e.g., suicide), and health care worker psycho-social impacts (stress, fatigue, and burn out) which may need further evaluation when applying the results of this model.

One important limitation of compartmental models, as used in this paper, are less suitable to model household infection dynamics since homes have limited numbers of individuals and once infected, they are removed from the ongoing epidemic. This modeling weakness is difficult to incorporate by compartmental models in general, so incidence may be overestimated. On the other hand, since household contact rates tend to increase during lockdowns the resulting incidence might also increase, counter-balancing this issue; which we did not incorporate in this model trying to overcome some of the inherent limitations to this type of compartmental modeling of household transmission. Also, all timing scenarios in this paper are subjected to the same limitation and so the qualitative rather than quantitative findings of the paper are to be considered.

Finally, estimation of the pandemic peak by individual countries at the start of a pandemic with limited epidemiological case data remains a significant challenge for public health officials. Accurately timing lockdowns to achieve a “tunneling effect” is vital to maximize its benefits. Our results endorse that hypothetical lockdown scenarios for representative countries (Canada, China, Mexico, and Niger) spanning a continuum of increasing rates of social contact can all benefit from well-timed lockdown interventions.

## Data Availability

N/A

## Acknowledgements

M.A. gratefully acknowledge the assistance and support of Kuwait University and the Kuwait Foundation for the Advancement of Sciences (KFAS) grant number “CORONA PROP 46” to complete this research.

## Authors Contribution

TO, MGT, SE and MA conceived conception and designed the model.

TO worked on the mathematical model establishment, simulation, coding and analytical work.

TO, WA, MGT, and MA contributed to the acquisition of data.

TO, MGT, MA, KV, JCL, WQA and MA contributed to analysis and interpretation of outputs.

TO, MGT, JCM, KV, SE, WQA, JCL and MA worked on reviewing different drafts of the manuscript.

TO, MGT, JCM, KV, SE, WQA, JCL and MA contributed to the final revision.

All authors reviewed the final manuscript.

## References

1. Majumder, M and Mandl, K. Early Transmissibility Assessment of a Novel Coronavirus in Wuhan, China (January 26, 2020). Available at SSRN: https://ssrn.com/abstract=3524675 or http://dx.doi.org/10.2139/ssrn.3524675

2. World Health Organization. Statement on novel coronavirus in Thailand. Jan 13, 2020. Available at https://www.who.int/news-room/detail/13-01-2020-who-statement-on-novel-coronavirus-in-thailand

3. World Health Organization. Statement on the second meeting of the International Health Regulations (2005) Emergency Committee regarding the outbreak of novel coronavirus (2019-nCoV). Jan 30, 2020. Available at https://www.who.int/news-room/detail/30-01-2020-statement-on-the-second-meeting-of-the-international-health-regulations-(2005)-emergency-committee-regarding-the-outbreak-of-novel-coronavirus-(2019-ncov) (2020).

4. Pan A, Liu L, Wang C, et al. Association of Public Health Interventions with the Epidemiology of the COVID-19 Outbreak in Wuhan, China. JAMA. 323(19):1915–1923 (2020). doi:10.1001/jama.2020.6130

5. Schuchat A. Public Health Response to the Initiation and Spread of Pandemic COVID-19 in the United States, February 24–April 21, 2020. MMWR Morb Mortal Wkly Rep. 69:551–556 (2020). DOI: http://dx.doi.org/10.15585/mmwr.mm6918e2externalicon

6. Porcheddu, R., Serra, C., Kelvin, D., Kelvin, N. & Rubino, S. Similarity in Case Fatality Rates (CFR) of COVID-19/SARS-COV-2 in Italy and China. J Infect Dev Ctries. 14(2), 125–128 (2020).

7. Flaxman, S. et al.. Estimating the effects of non-pharmaceutical interventions on COVID-19 in Europe. Nature. 10.1038/s41586-020-2405-7 (2020).

8. Güner, R., Hasanoğlu, I. & Aktaş, F. COVID-19: Prevention and control measures in community. Turk J Med Sci. 50(SI-1), 571–577 (2020).

9. Ebrahim, S. H., Ahmed, Q. A., Gozzer, E., Schlagenhauf, P. & Memish Z. A. Covid-19 and community mitigation strategies in a pandemic. BMJ. 368, m1066 (2020).

10. Hou, C. et al.. The effectiveness of quarantine of Wuhan city against the Corona Virus Disease 2019 (COVID-19): A well-mixed SEIR model analysis. J Med Virol. 92(7), 841–848 (2020).

11. Wilder-Smith, A. & Freedman, D. O. Isolation, quarantine, social distancing and community containment: pivotal role for old-style public health measures in the novel coronavirus (2019-nCoV) outbreak. J Travel Med. 27(2), taaa020 (2020).

12. Allen, L. J. & van den Driessche, P. Relations between deterministic and stochastic thresholds for disease extinction in continuous- and discrete-time infectious disease models. Math Biosci. 243(1), 99–108 (2013).

13. Prem, K., Cook, A. R. & Jit, M. Projecting social contact matrices in 152 countries using contact surveys and demographic data. PLoS Comput Biol. 13(9), e1005697 (2017).

14. Mossong, J. et al.. Social contacts and mixing patterns relevant to the spread of infectious diseases. PLoS Med. 5(3), e74 (2008).

15. Pellis, L., Cauchemez, S., Ferguson, N.M. et al.. Systematic selection between age and household structure for models aimed at emerging epidemic predictions. Nat Commun 11(906), (2020). https://doi.org/10.1038/s41467-019-14229-4

16. United Nations. 2019. Data Query. Department of Economic and Social Affairs Population Dynamics. https://population.un.org/wpp/DataQuery/ (2020).

17. Yang, C. & Gillespie, D. T. Efficient step size selection for the tau-leaping simulation method. J. Chem. Phys. 124, 044109 (2006).

18. Allen, L. J. S. & Lahodny, G. E. Jr. Extinction thresholds in deterministic and stochastic epidemic models. J Biol Dyn. 6(2), 590–611 (2012).

19. Tang, B. et al.. Estimation of the transmission risk of the 2019-nCoV and its implication for public health interventions. J Clin Med. 9(2), 462 (2020).

20. Munasinghe, M. Is environmental degradation an inevitable consequence of economic growth: tunneling through the environmental Kuznets curve. Ecol Econ. 29(1), 89–109 (1999).

21. Greenwood, P. E. & Gordillo, L. F. Stochastic epidemic modeling. In Mathematical and Statistical Estimation. 366, Approaches in Epidemiology. 31–52. (Springer, 2009).

22. Carcione, J. M., Santos, J. E., Bagaini, C. & Ba, J. A simulation of a COVID-19 epidemic based on a deterministic SEIR model. Front Public Health. 8, 230 (2020).

23. Reno, C. et al.. Forecasting COVID-19-associated hospitalizations under different levels of social distancing in Lombardy and Emilia-Romagna, Northern Italy: results from an extended SEIR compartmental model. J Clin Med. 9(5), 1492 (2020).

24. Bausch, D. G. Precision physical distancing for COVID-19: an important tool in unlocking the lockdown. Am J Trop Med Hyg. 103(1), 22–24 (2020).

25. World Health Organization. Statement on the fourth meeting of the International Health Regulations (2005) Emergency Committee regarding the outbreak of coronavirus disease (COVID-19). Available at https://www.who.int/news-room/detail/01-08-2020-statement-on-the-fourth-meeting-of-the-international-health-regulations-(2005)-emergency-committee-regarding-the-outbreak-of-coronavirus-disease-(covid-19)

26. Ngonghala, C. N. et al. Mathematical assessment of the impact of non-pharmaceutical interventions on curtailing the 2019 novel Coronavirus. Math Biosci. 325, 108364 (2020).

27. Imai, N. et al.. Adoption and impact of non-pharmaceutical interventions for COVID-19. Wellcome Open Res. 5, 59 (2020).

28. Lau, H. et al.. The positive impact of lockdown in Wuhan on containing the COVID-19 outbreak in China. J Travel Med. 27(3), taaa037 (2020).

29. Ji, T. et al.. Lockdown contained the spread of 2019 novel coronavirus disease in Huangshi city, China: Early epidemiological findings. Clin Infect Dis. ciaa390 (2020).

