## Supplementary Material for "Modeling the Effect of Lockdown Timing as a COVID-19 Control Measure in Countries with Differing Social Contacts"

*S1. Model Description*

A CTMC can capture the initial disease dynamics and accommodate the uncertainties involved in the disease transmission process. It is also recommended given the small number of cases in the beginning of the epidemics. We assume an initial random number of individuals in the E, A, and I compartment given by $e_0$, $a_0$, and $i_0$, respectively. The probability of transitions of the CTMC, $\boldsymbol{X}(t)$, from state $x$ to state $y$ (denoted by $x \to y$) in the interval $(t, t + dt]$ for very small $dt$ is given by

$$P(\boldsymbol{X}(t + dt) = y | \boldsymbol{X}(t) = x) = r_{xy}\, dt + o(dt)$$

with specific rates $r_{xy}$. The disease transitions in the underlying CTMC are occurring according to the descriptions and rates given in Table S1.

**Table S1. Description of modeled transitions and their rates of the CTMC between the SEAMHQRD-V compartments.**

| Transitions | Numbers in compartments that undergo changes while the rest compartments remain fixed | Rates ($\boldsymbol{r_{xy}}$) |
|---|---|---|
| Becoming Exposed (latent) | For $i = c, a,$ or $s$; $S_i \to S_i - 1$ and $E_i \to E_i + 1$ | $\beta_i S_i ( \sum_{j=c,a,s} \tilde{C}_{ij}^{v} V_j + \sum_{j=c,a,s} \tilde{C}_{ij} \frac{A_j + M_j}{N_j} )$ |
| Becoming asymptomatic | For $i = c, a,$ or $s$; $E_i \to E_i - 1$ and $A_i \to A_i + 1$ | $\alpha(1-p)E_i$ |
| Becoming symptomatic and mild | For $i = c, a,$ or $s$; $E_i \to E_i - 1$ and $M_i \to M_i + 1$ | $\alpha\, p\, E_i$ |
| Becoming symptomatic and severe | For $i = c, a,$ or $s$; $M_i \to M_i - 1$ and $H_i \to H_i + 1$ | $\gamma_M M_i$ |
| Getting quarantined (isolated) | For $i = c, a,$ or $s$; $M_i \to M_i - 1$ and $Q_i \to Q_i + 1$ | $q M_i$ |
| Becoming severe and hospitalized | For $i = c, a,$ or $s$; $Q_i \to Q_i - 1$ and $H_i \to H_i + 1$ | $\gamma_Q Q_i$ |
| Recovery of asymptomatic | For $i = c, a,$ or $s$; $A_i \to A_i - 1$ and $R_i \to R_i + 1$ | $\mu_A A_i$ |
| Recovery of mildly infected | For $i = c, a,$ or $s$; $M_i \to M_i - 1$ and $R_i \to R_i + 1$ | $\mu_M M_i$ |
| Recovery of severely infected | For $i = c, a,$ or $s$; $H_i \to H_i - 1$ and $R_i \to R_i + 1$ | $\mu_H H_i$ |
| Recovery of quarantined children, adults and seniors | For $i = c, a,$ or $s$; $Q_i \to Q_i - 1$ and $R_i \to R_i + 1$ | $\mu_Q Q_i$ |
| Disease-specific death of children, adults and seniors | For $i = c, a,$ or $s$; $H_i \to H_i - 1$ and $D_i \to D_i + 1$ | $\sigma_i H_i$ |
| Environmental contamination by infected children, adults and seniors | For $i = c, a,$ or $s$; $V_i \to V_i + 1$ | $\tilde{\omega}_{A,i} A_i + \tilde{\omega}_{M,i} M_i$ |
| Environmental contamination by infected children, adults and seniors | For $i = c, a,$ or $s$; $V_i \to V_i - 1$ | $\rho V_i$ |

We use five social-contact matrices $\boldsymbol{C}^k$ for $k = sc$ (school), $h$ (household), $w$ (work), $o$ (other), and $v$ (environment). defines the contact rates between the three age groups

$$\boldsymbol{C}^k = \begin{pmatrix} C_{cc}^k & C_{ca}^k & C_{cs}^k \\ C_{ac}^k & C_{aa}^k & C_{as}^k \\ C_{sc}^k & C_{sa}^k & C_{ss}^k \end{pmatrix} \tag{1}$$

A socially altered contact matrix $\widetilde{\boldsymbol{C}}$ defines the contact rates between the three age groups with social distances and lockdown, where the column vector. The entries of $\widetilde{\boldsymbol{C}}$ are defined by

$$\tilde{C}_{ij} = C_{ij}^h + \sum_{k=sc,w,o} C_{ij}^k \left(1 - \tilde{p}_j^k(t)\right)\left(1 - \tilde{p}_i^k(t)\right)$$

and

$$\tilde{C}_{ij}^v = C_{ij}^v\left(1 - \tilde{p}_i^v(t)\right)$$

for $i$ and $j = c, a, s$. The vector $\left(p_c^k, p_a^k, p_s^k\right)$ defines the proportion of those practicing social distances in the three age groups at the different location types $k = sc, w, o, v$, and so degree of adherence to social distancing. The parameter $\tilde{p}_i^k$ is the government imposed closures and enforced stay-home which only takes values $\tilde{p}_i$ based on a policy turning points $t_{l,i}$ in a way that $\tilde{p}_i^k(t) = p_i^k\ I\left(t_{1,i} \geq t \geq t_{0,i}\right)$.

The environmental transmission has the component at time t *is* $\mathrm{V_j}$ for $j = c, a,$ and $s$, where

$$\widetilde{\omega}_{\mathrm{A,i}}(t) = \omega_{\mathrm{A}}\left(1 - \tilde{p}_i^v(t)\right)$$

$$\widetilde{\omega}_{\mathrm{M,i}}(t) = \omega_{\mathrm{M}}\left(1 - \tilde{p}_i^v(t)\right)$$

where $\omega_{\mathrm{A}}$ $(\omega_{\mathrm{M}})$ is the number of individuals equivalent to environmental contamination/deposit made by asymptomatic (mildly infected) individual per place.

*S2. The Basic Reproduction Number* $\mathrm{R_0}$ *and Probability of Extinction*

In this section, for brevity, we use subscripts 1, 2, and 3 for children, adults, and seniors age groups, respectively. We follow Allen [1] by approximating the beginning of an epidemic by a multi-type branching process for the types $(\mathrm{A_1, M_1, V_1, A_2, M_2, V_2, A_3, M_3, V_3})$, with extremely large initial number of susceptible in each age group to be almost equal to the sizes of their corresponding sub-populations. We use the offspring generating functions of the nine types $(\mathrm{A_1, M_1, V_1, A_2, M_2, V_2, A_3, M_3, V_3})$ that results in infections. The offspring rates for types $\mathrm{A_i, M_i, V_i}$ by types $\mathrm{A_j, M_j, V_j}$ for $\mathrm{i, j} = 1,2,3$ are summarized by the following table.

**Table S2. Offspring probabilities of column types by row types.**

| | $\mathrm{A_i}$ | $\mathrm{M_i}$ | $\mathrm{V_i}$ | *vanishes* |
|---|---|---|---|---|
| $\mathrm{A_j}$ | $\beta_i N_i \frac{\tilde{C}_{ij}}{N_j}(1-p)$ | $\beta_i N_i \frac{\tilde{C}_{ij}}{N_j}\ p$ | $\widetilde{\omega}_{\mathrm{A,j}}\delta_{\mathrm{ij}}$ | $\mu_{\mathrm{A}}$ |
| $\mathrm{M_j}$ | $\beta_i N_i \frac{\tilde{C}_{ij}}{N_j}(1-p)$ | $\beta_i N_i \frac{\tilde{C}_{ij}}{N_j}\ p$ | $\widetilde{\omega}_{\mathrm{M,j}}\delta_{\mathrm{ij}}$ | $q + \mu_{\mathrm{M}} + \gamma_M$ |
| $\mathrm{V_j}$ | $\beta_i N_i \widetilde{C}_{ij}^v(1-p)$ | $\beta_i N_i \widetilde{C}_{ij}^v\ p$ | 0 | $\rho$ |

The offspring probability generating functions with arguments $(u_{1,1}, u_{1,2}, u_{1,3}, u_{2,1}, u_{2,2}, u_{2,3}, u_{3,1}, u_{3,2}, u_{3,3})$ for the types $(\mathrm{A}_1, \mathrm{M}_1, \mathrm{V}_1, \mathrm{A}_2, \mathrm{M}_2, \mathrm{V}_2, \mathrm{A}_3, \mathrm{M}_3, \mathrm{V}_3)$ are then given by

$$f_{A_j}(u_{1,1}, u_{1,2}, u_{1,3}, u_{2,1}, u_{2,2}, u_{2,3}, u_{3,1}, u_{3,2}, u_{3,3}) = \frac{u_{j,1}\left[\sum_{i=1}^{3} \beta_i N_i \frac{\tilde{C}_{ij}}{N_j}(1-p)\, u_{i,1} + \sum_{i=1}^{3} \beta_i N_i \frac{\tilde{C}_{ij}}{N_j} p\, u_{i,2} + \tilde{\omega}_{\mathrm{A,j}} u_{j,3}\right] + \mu_{\mathrm{A}}}{d_{1,j}},$$

$$f_{M_j}(u_{1,1}, u_{1,2}, u_{1,3}, u_{2,1}, u_{2,2}, u_{2,3}, u_{3,1}, u_{3,2}, u_{3,3}) = \frac{u_{j,2}\left[\sum_{i=1}^{3} \beta_i N_i \frac{\tilde{C}_{ij}}{N_j}(1-p)\, u_{i,1} + \sum_{i=1}^{3} \beta_i N_i \frac{\tilde{C}_{ij}}{N_j} p\, u_{i,2} + \tilde{\omega}_{\mathrm{M,j}} u_{j,3}\right] + q + \mu_{\mathrm{M}} + \gamma_M}{d_{2,j}},$$

and

$$f_{V_j}(u_{1,1}, u_{1,2}, u_{1,3}, u_{2,1}, u_{2,2}, u_{2,3}, u_{3,1}, u_{3,2}, u_{3,3}) = \frac{u_{j,3}\left[\sum_{i=1}^{3} \beta_i N_i \tilde{C}_{ij}^{v}(1-p)\, u_{i,1} + \sum_{i=1}^{3} \beta_i N_i \tilde{C}_{ij}^{v} p\, u_{i,2}\right] + \rho}{d_{3,j}},$$

where

$$d_{1,j} = \sum_{i=1}^{3} \beta_i N_i \frac{\tilde{C}_{ij}}{N_j} + \tilde{\omega}_{\mathrm{A,j}} + \mu_{\mathrm{A}},$$

$$d_{2,j} = \sum_{i=1}^{3} \beta_i N_i \frac{\tilde{C}_{ij}}{N_j} + \tilde{\omega}_{\mathrm{M,j}} + q + \mu_{\mathrm{M}} + \gamma_M,$$

and

$$d_{3,j} = \sum_{i=1}^{3} \beta_i N_i \tilde{C}_{ij}^{v} + \rho.$$

for $j = 1,2,3$. The generating functions are not simple. The mean offspring matrix $\mathbf{M}$ is a $9 \times 9$ matrix whose elements could be derived by differentiating the generating functions with respect to $u_{j,k}$ and then substiting for all $u_{j,k}$'s by one. With some work, we can reach that

$$(\mathbf{M} - \mathbf{I_9})\boldsymbol{\Lambda} = \mathbf{B} \otimes \widetilde{\mathbf{P}} - \mathbf{I_3} \otimes \mathbf{V}$$

where $\mathbf{I_n}$ is the $n \times n$ identity matrix and $\otimes$ is the Kronecker product. The matrix $\mathbf{M}$ is irreducible. With the assumption that $\tilde{C}_{ij}^{v} = r\frac{\tilde{C}_{ij}}{N_j}$ and $\tilde{p}_i^{v}$ *is not dependent on the age group*, the matrices are

$$\mathbf{B} = \begin{pmatrix} \beta_1 N_1 \frac{\tilde{C}_{11}}{N_1} & \beta_1 N_1 \frac{\tilde{C}_{12}}{N_2} & \beta_1 N_1 \frac{\tilde{C}_{13}}{N_3} \\ \beta_2 N_2 \frac{\tilde{C}_{21}}{N_1} & \beta_2 N_2 \frac{\tilde{C}_{22}}{N_2} & \beta_2 N_2 \frac{\tilde{C}_{23}}{N_3} \\ \beta_3 N_3 \frac{\tilde{C}_{31}}{N_1} & \beta_3 N_3 \frac{\tilde{C}_{32}}{N_2} & \beta_3 N_3 \frac{\tilde{C}_{33}}{N_3} \end{pmatrix} = \begin{pmatrix} \beta_1 N_1 & 0 & 0 \\ 0 & \beta_2 N_2 & 0 \\ 0 & 0 & \beta_3 N_3 \end{pmatrix} \tilde{\boldsymbol{C}} \begin{pmatrix} \frac{1}{N_1} & 0 & 0 \\ 0 & \frac{1}{N_2} & 0 \\ 0 & 0 & \frac{1}{N_3} \end{pmatrix}$$

$$\widetilde{\mathbf{P}} = \begin{pmatrix} 1-p & 1-p & r(1-p) \\ p & p & rp \\ 0 & 0 & 0 \end{pmatrix}$$

$$\mathbf{V} = \begin{pmatrix} \mu_A & 0 & 0 \\ 0 & q+\mu_M+\gamma_M & 0 \\ -\widetilde{\omega}_A & -\widetilde{\omega}_M & \rho \end{pmatrix}$$

and $\mathbf{\Lambda}$ is the diagonal matrix whose entries are given by $d_{1,1}, d_{2,1}, d_{3,1}, d_{1,2}, d_{2,2}, d_{3,2}, d_{1,3}, d_{2,3}, d_{3,3}$. Thus, by Theorem A1 in [2] it is easy to find that

$$R_0 = \boldsymbol{\rho}(\mathbf{B} \otimes \widetilde{\mathbf{P}}\mathbf{V}^{-1}) = \boldsymbol{\rho}(\mathbf{B})\boldsymbol{\rho}(\widetilde{\mathbf{P}}\mathbf{V}^{-1})$$

where $\boldsymbol{\rho}$ is the spectral radius of the matrix and

$$\mathbf{V}^{-1} = \begin{pmatrix} \mathbf{1}/\mu_A & \mathbf{0} & \mathbf{0} \\ \mathbf{0} & 1/(q+\mu_M+\gamma_M) & \mathbf{0} \\ \widetilde{\omega}_A/\rho\mu_A & \widetilde{\omega}_M/\rho(q+\mu_M+\gamma_M) & 1/\rho \end{pmatrix}$$

*Also,*

$$\boldsymbol{\rho}(\widetilde{\mathbf{P}}\mathbf{V}^{-1}) = (1-p)\frac{1 + r\,\widetilde{\omega}_A/\rho}{\mu_A} + p\,\frac{1 + r\,\widetilde{\omega}_M/\rho}{q+\mu_M+\gamma_M}$$

Therefore, if

$$R_0 = \boldsymbol{\rho}(\mathbf{B})\left[(1-p)\frac{1 + r\,\widetilde{\omega}_A/\rho}{\mu_A} + p\,\frac{1 + r\,\widetilde{\omega}_M/\rho}{q+\mu_M+\gamma_M}\right]$$

is less than one then the epidemic dies out with probability one. If $R_0$ is more than one then there exists unique fixed points $0 < q_{1,1}, q_{1,2}, q_{1,3}, q_{2,1}, q_{2,2}, q_{2,3}, q_{3,1}, q_{3,2}, q_{3,3} < 1$ that solve the system of equations

$$f_{A_j}(q_{1,1}, q_{1,2}, q_{1,3}, q_{2,1}, q_{2,2}, q_{2,3}, q_{3,1}, q_{3,2}, q_{3,3}) = q_{j,1},$$

$$f_{M_j}(q_{1,1}, q_{1,2}, q_{1,3}, q_{2,1}, q_{2,2}, q_{2,3}, q_{3,1}, q_{3,2}, q_{3,3}) = q_{j,2},$$

and

$$f_{V_j}(q_{1,1}, q_{1,2}, q_{1,3}, q_{2,1}, q_{2,2}, q_{2,3}, q_{3,1}, q_{3,2}, q_{3,3}) = q_{j,3}$$

for $j = 1,2,3$. Thus the probability of extinction is of an epidemic that starts with $(A_1(0), M_1(0), V_1(0), A_2(0), M_2(0), V_2(0), A_3(0), M_3(0), V_3(0)) = (a_1, m_1, v_1, a_2, m_2, v_2, a_3, m_3, v_3)$

is given by

$$\mathbf{Q} = \prod_{i=1}^{3}\left({q_{i,1}}^{a_i}.\,{q_{i,2}}^{m_i}.\,{q_{i,3}}^{v_i}\right)$$

*S3. Countries Categorization*

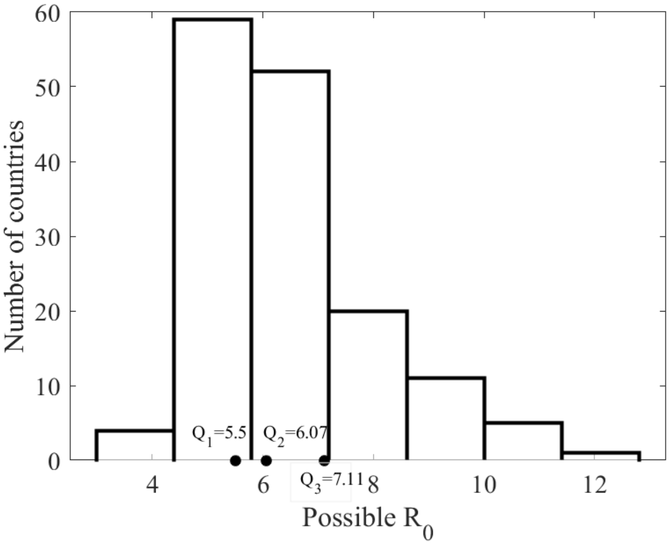


**Figure S1.** Histogram of possible $R_0$ from 152 countries [3] calculated at $\beta = 3.5\%$, see the table below. Countries were split into 4 categories based on the quartiles of all of the 152 values of $R_0$, which come to be $Q_1 = 5.5$, $Q_2 = 6.07$, and $Q_3 = 7.11$. The basic reproduction numbers of the four selected countries are: 4.86 for Canada, 5.75 for China, 6.12 for Mexico, and 12.30 for Niger. The possible $R_0$ might have extreme values but that is only a reflection of the effect of contact rates and household age structure in case of no control measures.

*S4. Model Parameterization*

**Table S3. A list of parameters of the CTMC between the SEAMHQRD-V compartments.**

| Type | Parameters or variables | Description | Base value | Range | Source Reference |
|---|---|---|---|---|---|
| **Demographic** | $N_c$ | Children's population (0-18 y) size | Based on country | 2019 Population age distribution | [4] |
| | $N_a$ | Adults' population (19-64 y) size | Based on country | 2019 Population age distribution | [4] |
| | $N_s$ | Seniors' population (65+ y) size | Based on country | 2019 Population age distribution | [4] |
| | $(m_{0,c}, m_{0,a}, n$ | Initial number of mildly infected and symptomatic individuals (children, adults, seniors) | (0,1,0) | 0 or 1 | Assumption |
| **Disease-specific parameters** | $\beta_c$ | Children infection probability upon contact with an | 1.3 % | | [5] |

| | | | | | |
|---|---|---|---|---|---|
| | | infectious individual | | | |
| | $\beta_a$ | Adults infection probability upon contact with an infectious individual | 3.5% | | [5] |
| | $\beta_s$ | Senior infection probability upon contact with an infectious individual | 3.5% | | [5] |
| | α | Rate of removal from exposed compartment (per day) | 1/5days = 0.2 per day | 3 to 7 days incubation time = 0.33-0.142 per day (range)<br><br>[the longest time from infection to symptoms was 12.5 days (95% CI, 9.2 to 18)] | [6] |
| | $p$ | Probability of showing symptoms among those exiting the exposed compartment | 0.821 | (estimated 17.9% were asymptomatic)<br><br>Range: 0.5-0.9 | [7] |
| | $\gamma_M$ | Rate of progression from mild to severe infection | 1/8= 0.125<br><br>8 days<br><br>9 days | [8] Dypsnea (severe symptoms) 8 days after illness onset, range: 5–13 days.<br><br>1/5-1/13<br>= 0.2-0.076 per day<br><br>[9] "the mean time from illness onset to hospital admission with pneumonia was 9 days" | [8]<br><br>[9] |
| | $\gamma_Q$ | Rate of progression from quarantine to severe infection | | | |
| | $\mu_M$ ($\mu_A$) | Recovery rate for mildly infected (and asymptomatic) | 0.05 (20 days median)<br><br>14-37 days* | [10] reported that viral shedding continued for a median of 20 days (maximum 37 days) in survivors and until death in non-survivors.<br><br>[11] "33% patients of the 99 infected have been discharged within 5-20 days". If we assume that viral shedding reflects the existence of disease then assume 14-37 days as indicated by the descriptive cases. | [10]<br><br>[11] |

| | | | | | |
|---|---|---|---|---|---|
| | $\mu_H$ | Recovery rate for severely infected | 30 days<br><br>Minimum 20 days (range 8-37 days) (severe cases shedding virus) | Depends on severity. Best guess based on similarity to SARS<br>Assume severe under quarantine are 100% contained<br><br>"Duration of viral shedding ranged between 8 and 37 days. The median duration of viral shedding was 20·0 days (IQR 17·0–24·0) in survivors, but continued until death in fatal cases." | [10] |
| | $\sigma_i$; for $i = c, a, s$ | Disease-specific death rate for children adults and seniors | -For children: 0/31 days<br><br>-For adults: <1% x 4,226 = < 42.26 over 31 days<br><br>among those aged 20-54 and 1-3% for those aged 55-64<br><br>-For seniors: 3-11% x 4,226 =<br><br>126.78 - 464.86 Over 31 days<br><br>among persons aged 65-84 and 10-27%x 4226 for persons aged >=85 | [12] CDC updates of patients with coronavirus<br><br>[13] Italy specific death rate is 7.2% (1625 deaths/22 512 cases, from Feb 20 to March 17. Not parsed by age. | [12]<br><br>[13] |

| | | | | | |
|---|---|---|---|---|---|
| | | | 426-1,141.02 over 31 days | | |
| **Social mixing and contact** | $C$ and $C^H$ | Social and household contact matrices (rates per day) | See Equation (1)<br><br>Singapore SARS 6.2% [95% confidence interval 3.9% to 8.6%]<br><br>China SARS 4.6%<br><br>Canada SARS 10.%<br><br>Hong Kong SARS 8% | Secondary household transmissions were low. Estimated from SARS reports.<br><br>[14] Singapore: The secondary household attack rate was thus low (6.2% [95% confidence interval 3.9% to 8.6%]). These findings are in contrast to the high attack rate seen in the healthcare setting (6). One possible explanation for this difference is the phase of the illness. SARS case-patients in the household tend to be in the early phase of illness whereas SARS case-patients in the healthcare settings tend to be in the later phase.<br><br>[15] China: Efficiency of quarantine during SARS<br><br>[16] Hong Kong: Secondary household transmissions<br><br>[17] Canada: Household-member secondary-attack rate we found, 10.2%. One of the most important factors for household transmission was duration of exposure in the home. We found a linear association between the number of days the ill index case remained at home and the secondary attack rate. These findings are in contrast to the high attack rate seen in the healthcare setting. One possible explanation for this difference is the phase of the illness. SARS case-patients in the household tend to be in the early phase of illness whereas SARS case-patients in the healthcare settings tend to be in the later phase. | [14]<br><br>[15]<br><br>[16]<br><br>[17] |

| | | | | | |
|---|---|---|---|---|---|
| | $p_i(t)$ for $i = c, a, s$ | Proportion of individuals practicing social distancing as a function in time | See Equation (3) $b_{0,i}, b_{1,i}, b_{2,i}$ $\tilde{p}i,$ $t_{0,i}$ $\tilde{p}_c = \tilde{p}_s = 1, \tilde{p}_a = .5$ | | |
| | $q$ | Probability of mildly infected going into self-quarantine | 0.9 | Assumption: Very few people will disregard public health officials.<br><br>First travel related patients and contact tracing by public health officials would place all identified people and contacts in to "home quarantine". Public health would seek 100% of contacts and first cases to be self-quarantined. | SARS expert estimate |
| **Environmental variables** | $C^V = r\ C^{All}$ | Contact rates with environment | R=1/6 | | |
| | $\omega_A$ | *Number of individuals equivalent to environmental contamination/deposit made by asymptomatic individual per place* | 1 | | |
| | $\omega_M$ | *Number of individuals equivalent to environmental contamination/deposit made by mildly infected individual per place* | 1 | | |
| | $\rho$ | *Natural removal rate of the environmental contamination* | 1/3 | | |
| | $K$ | *Cleaning rate of the environmental contamination* | 0 | | |

*S5. Additional Figures*

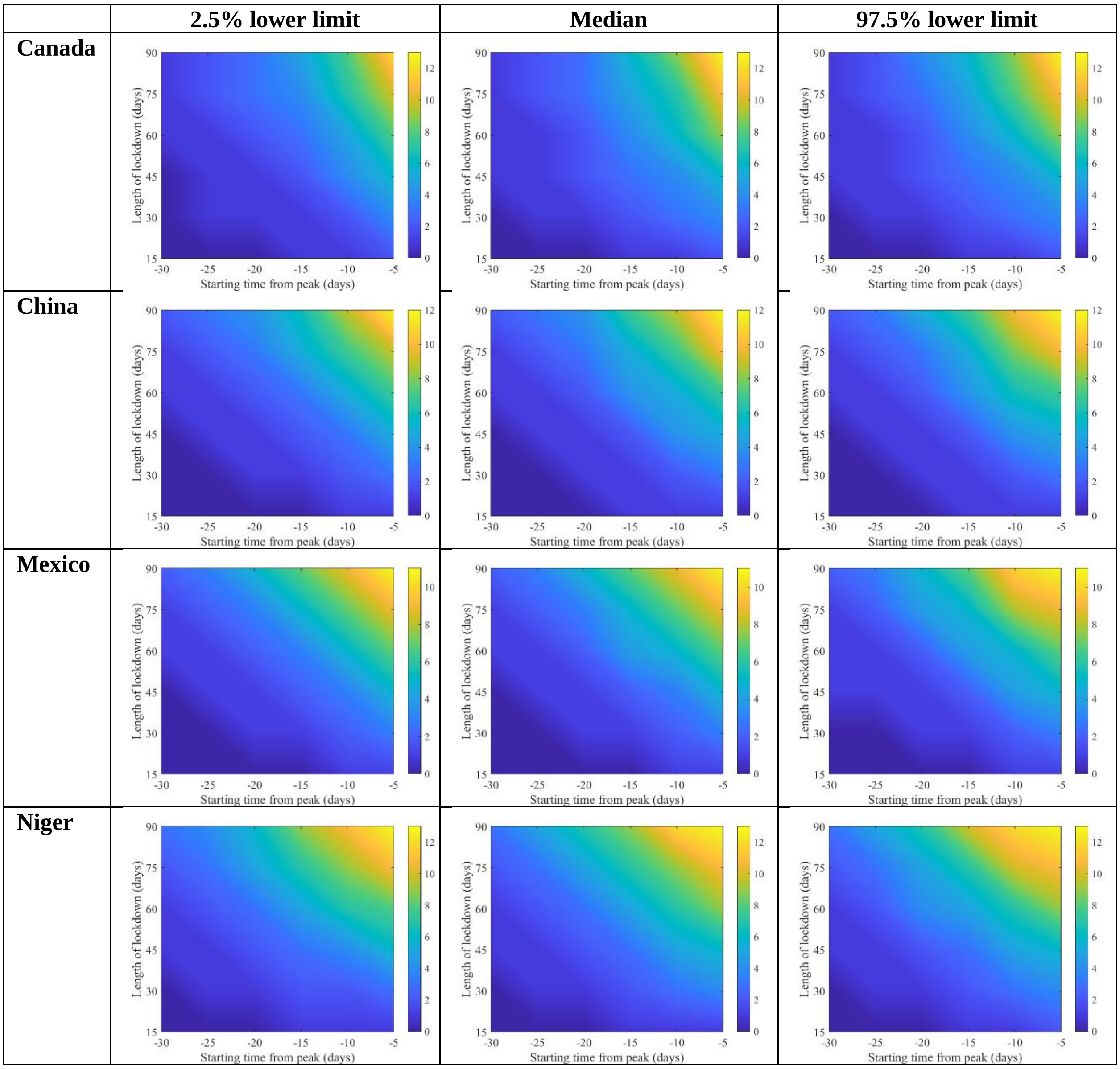


**Figure S2.** Median and 95% quartile interval of percentage relative reduction in attack rates (see Equation (1)) for (a) Canada, (b) China, (c) Mexico, and (d) Niger. They are calculated at $R_0 = 6.47$, with initially one adult mild infection. Bars to the right of the figures are percentages.

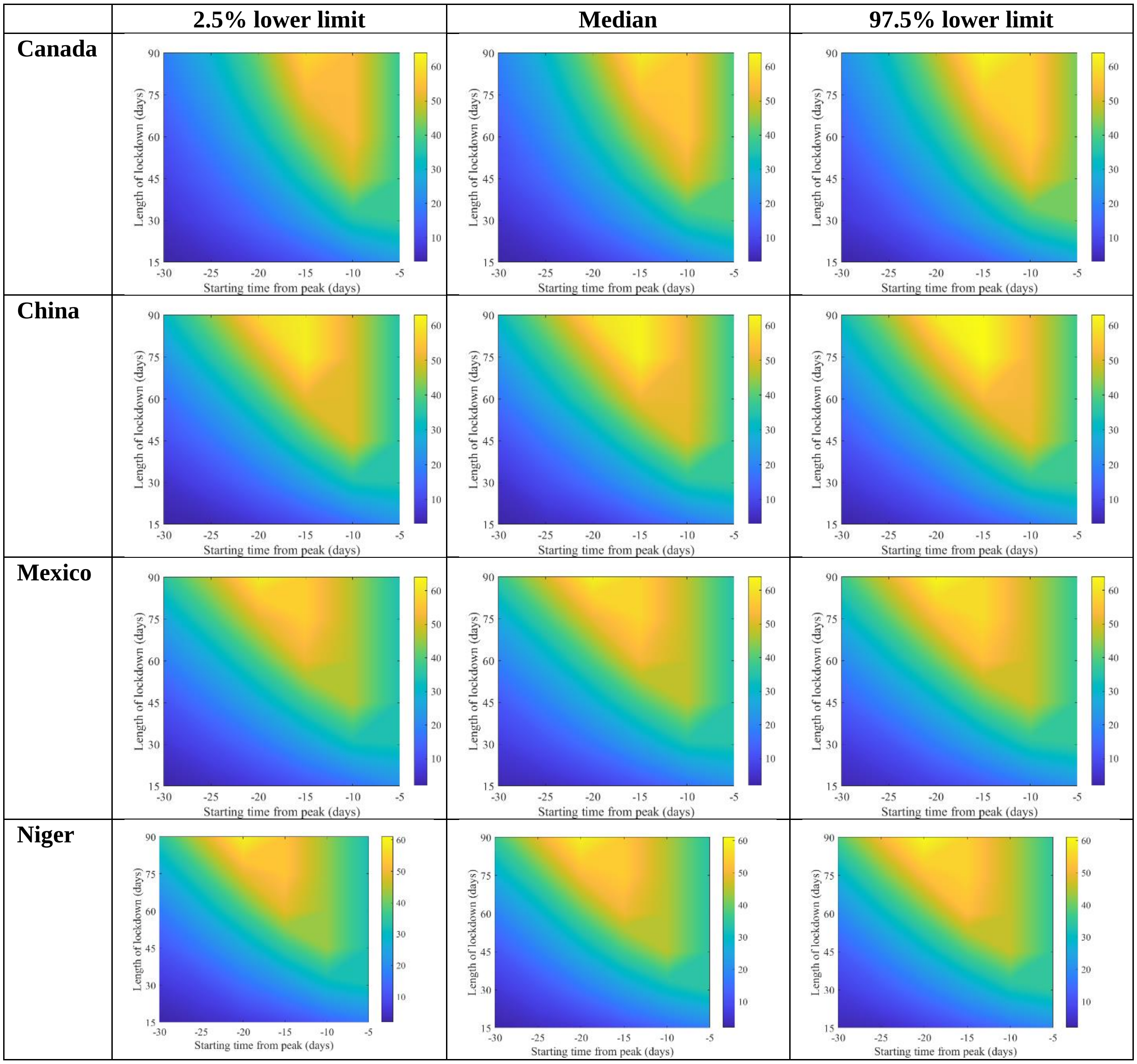


**Figure S3.** Median and 95% quartile interval of percentage relative reduction in hospitalization peak (see Equation (1)) for (a) Canada, (b) China, (c) Mexico, and (d) Niger. They are calculated at $R_0 = 6.47$, with initially one adult mild infection. Bars to the right of the figures are percentages.

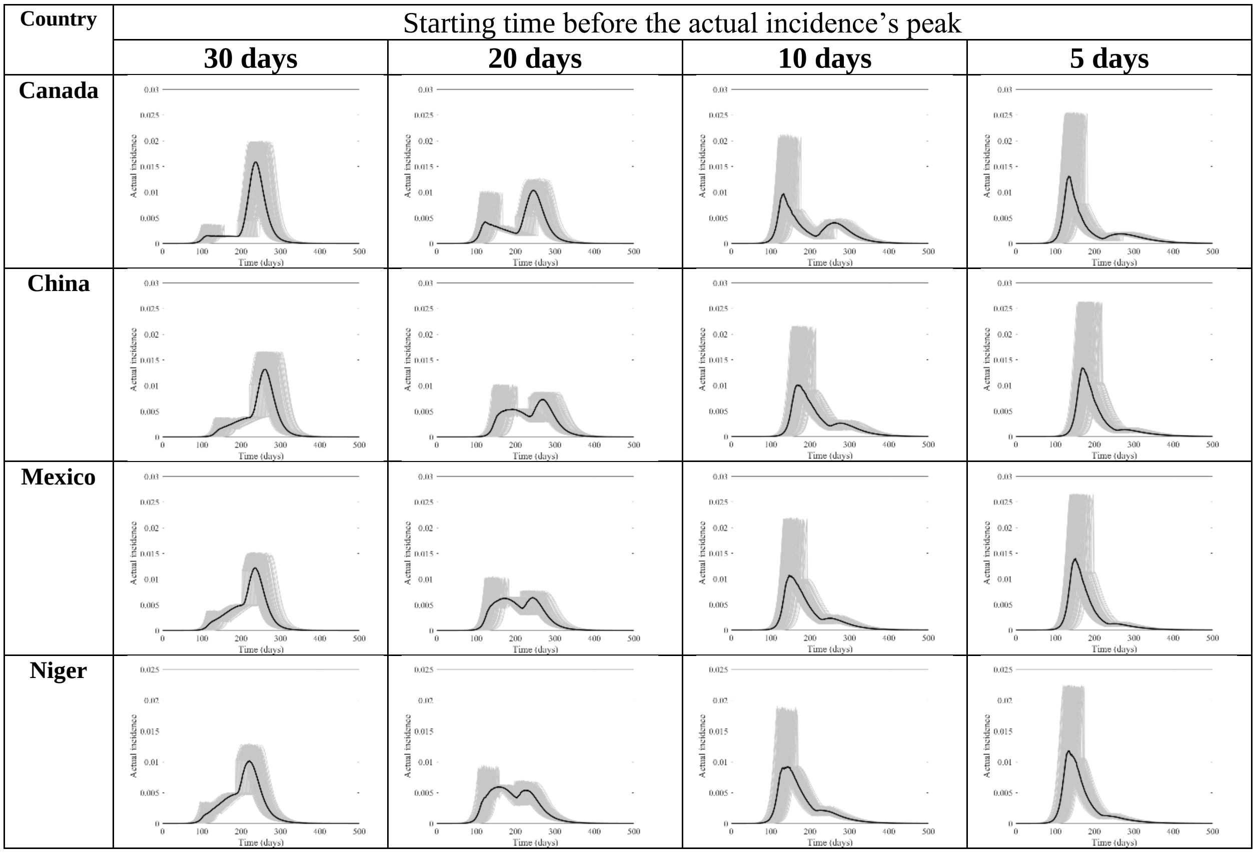


**Figure S4.** Actual incidence for Canada, China, Mexico, and Niger at four different times (days) of starting the lockdown before the peak and lasting for 90 days. They are calculated at $R_0 = 6.47$, with initially one adult mild infection. The grey curves are resulting from the stochastic model simulations and the black curve is the mean of those grey curves. They are all normalized by the population size.

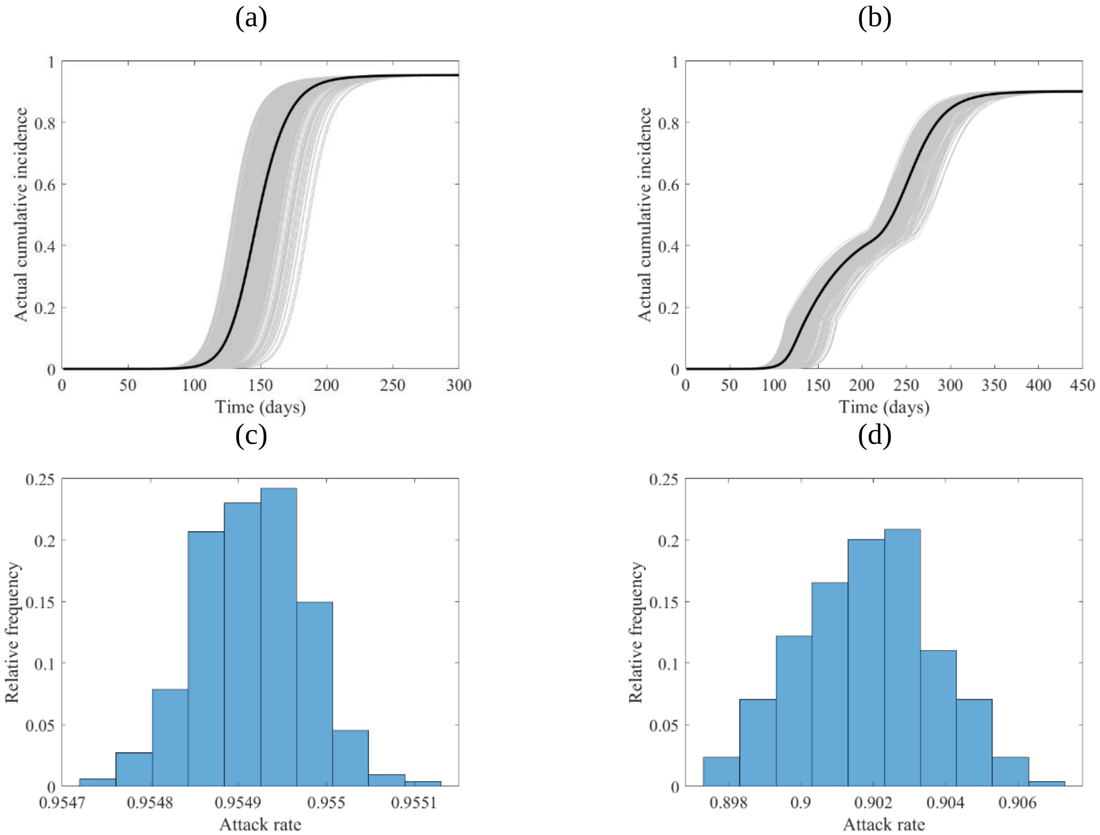


**Figure S5.** The course of the cumulative actual incidence (a) and (b), and relative frequency histogram of attack rate (c) and (d) in Canada with no control measure (left panel) and with stating lockdown (right panel) of 15 days before the peak and that lasts for 90 days. They are calculated at $R_0 = 6.47$, with initially one adult mild infection.

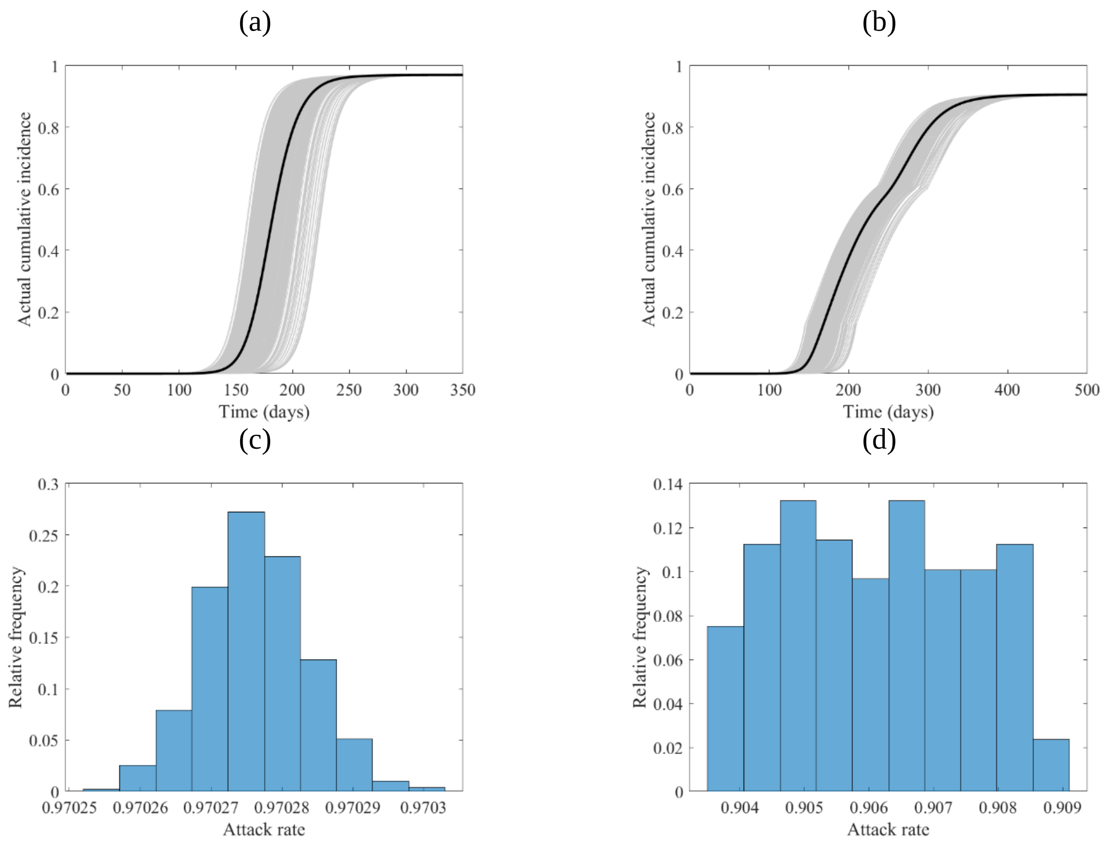


**Figure S6.** The course of the cumulative actual incidence (a) and (b), and relative frequency histogram of attack rate (c) and (d) in China with no control measure (left panel) and with stating lockdown (right panel) of 15 days before the peak and that lasts for 90 days. They are calculated at $R_0 = 6.47$, with initially one adult mild infection.

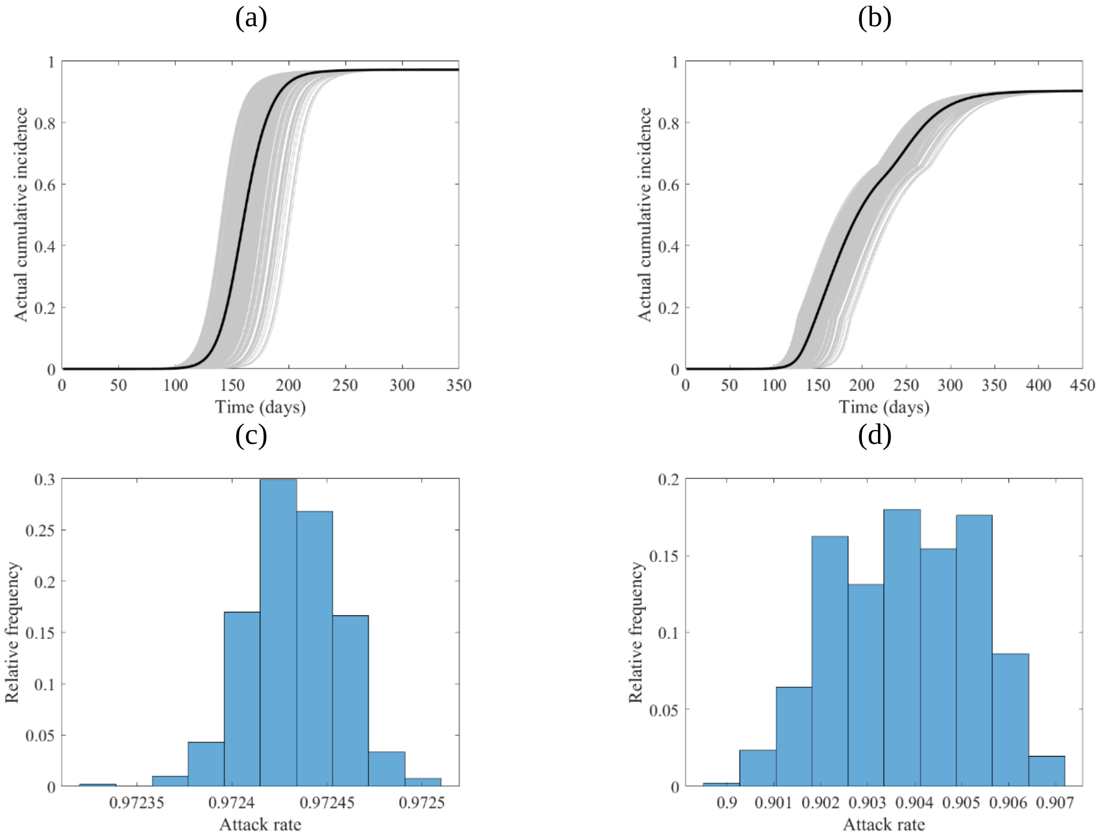


**Figure S7.** The course of the cumulative actual incidence (a) and (b), and relative frequency histogram of attack rate (c) and (d) in Mexico with no control measure (left panel) and with stating lockdown (right panel) of 15 days before the peak and that lasts for 90 days. They are calculated at $R_0 = 6.47$, with initially one adult mild infection.

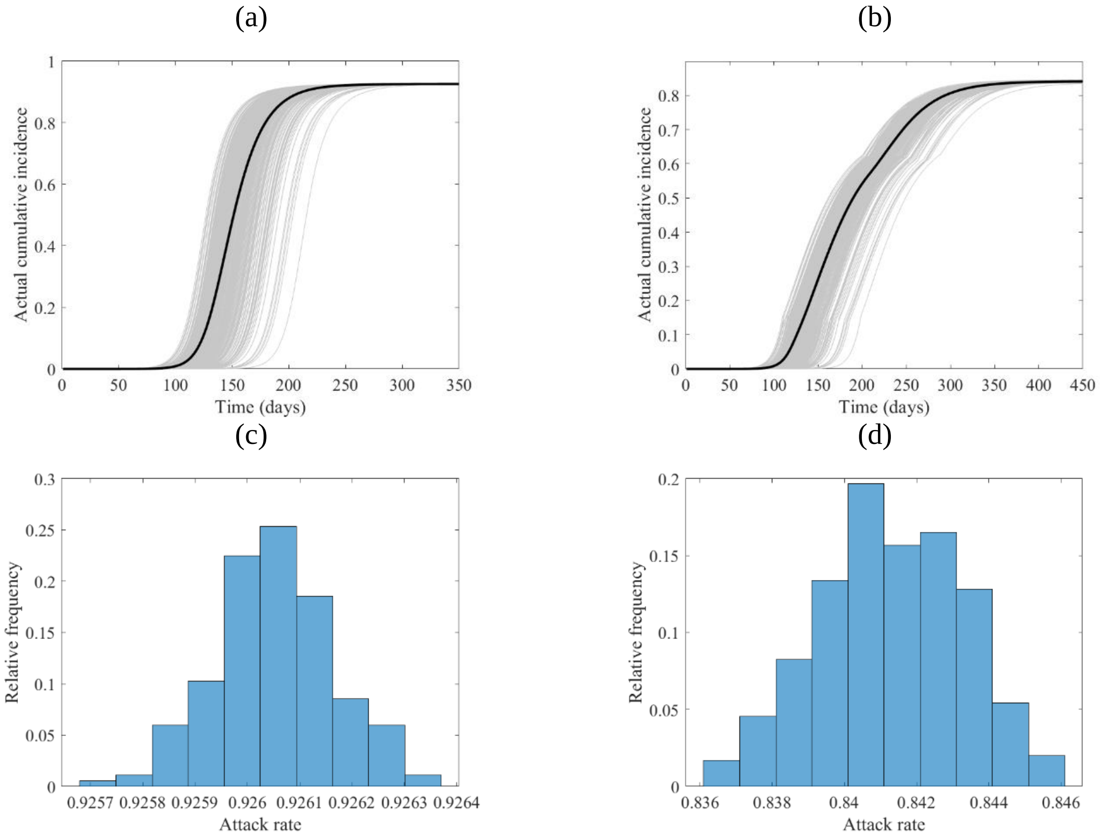


**Figure S8.** The course of the cumulative actual incidence (a) and (b), and relative frequency histogram of attack rate (c) and (d) in Niger with no control measure (left panel) and with stating lockdown (right panel) of 15 days before the peak and that lasts for 90 days. They are calculated at $R_0 = 6.47$, with initially one adult mild infection.

### References

[1] Allen, L. J. & van den Driessche, P. Relations between deterministic and stochastic thresholds for disease extinction in continuous- and discrete-time infectious disease models. *Math Biosci*. **243**(1), 99-108 (2013).

[2] Diekmann, O., Heesterbeek, J. A. & Roberts, M. G. The construction of next-generation matrices for compartmental epidemic models. *J R Soc Interface*. **47**, 873-885 (2010).

[3]. Prem, K., Cook, A. R. & Jit, M. Projecting social contact matrices in 152 countries using contact surveys and demographic data. *PLoS Comput Biol*. **13**(9), e1005697 (2017).

[4]. United Nations. 2019. Data Query. Department of Economic and Social Affairs Population Dynamics. https://population.un.org/wpp/DataQuery/ (2020).

[5]. Xu, Y. *et al*. Characteristics of pediatric SARS-CoV-2 infection and potential evidence for persistent fecal viral shedding. *Nat Med*. **26**(4), 502-505 (2020).

[6] Guo, Y. R. *et al*. The origin, transmission and clinical therapies on coronavirus disease 2019 (COVID-19) outbreak - an update on the status. *Mil Med Res*. **7**(1), 11 (2020).

[7] Mizumoto, K., Kagaya, K., Zarebski, A. & Chowell. G. Estimating the asymptomatic proportion of coronavirus disease 2019 (COVID-19) cases on board the Diamond Princess cruise ship, Yokohama, Japan, 2020. *Euro Surveill*. **25**(10), 2000180 (2020).

[8] Huang, C. *et al*. Clinical features of patients infected with 2019 novel coronavirus in Wuhan, China. *Lancet*. **395(**10223), 497-506 (2020).

[9] Guan, W. J. *et al.* China medical treatment expert group for Covid-19. Clinical characteristics of coronavirus disease 2019 in China. *N Engl J Med*. **382**(18), 1708-1720 (2020).

[10] Zhou *et al.* Clinical course and risk factors for mortality of adult inpatients with COVID-19 in Wuhan, China: a retrospective cohort study. *Lancet*. **395**(10229), 1038 (2020).

[11] Chen, N. *et al*. Epidemiological and clinical characteristics of 99 cases of 2019 novel coronavirus pneumonia in Wuhan, China: a descriptive study. *Lancet*. **395**(10223), 507-513 (2020).

[12] U.S. CDC. Severe outcomes among patients with coronavirus disease 2019 (COVID-19) - United States, February 12–March 16, 2020. *MMWR Morb Mortal Wkly Rep.* **69**, 343-346 (2020).

[13] Livingston, E. & Bucher, K. Coronavirus Disease 2019 (COVID-19) in Italy. *JAMA.* **323**(14), 1335 (2020).

[14] Goh, D. L. *et al.* Secondary household transmission of SARS, Singapore. *Emerg Infect Dis*. **10**(2), 232–234 (2004).

[15] U.S. CDC. Efficiency of quarantine during an epidemic of severe acute respiratory syndrome--Beijing, China, 2003. *MMWR Morb Mortal Wkly Rep*. **52**(43), 1037-40 (2003).

[16] Lau J. T., *et al*. Probable secondary infections in households of SARS patients in Hong Kong. *Emerg Infect Dis*. **10**(2), 235-43 (2004).

[17] Wilson-Clark, S. D. *et al.* Household transmission of SARS, 2003. *CMAJ*. **175**(10), 1219-1223 (2006).
